# Challenges at the *APOE* locus: A robust quality control approach for accurate *APOE* genotyping

**DOI:** 10.1101/2021.10.19.21265022

**Authors:** Michael E. Belloy, Sarah J. Eger, Yann Le Guen, Vincent Damotte, Shahzad Ahmad, M. Arfan Ikram, Alfredo Ramirez, Anthoula C. Tsolaki, Giacomina Rossi, Iris E. Jansen, Itziar de Rojas, Kayenat Parveen, Kristel Sleegers, Martin Ingelsson, Mikko Hiltunen, Najaf Amin, Ole Andreassen, Pascual Sánchez-Juan, Patrick Kehoe, Philippe Amouyel, Rebecca Sims, Ruth Frikke-Schmidt, Wiesje M. van der Flier, Jean-Charles Lambert, for the European Alzheimer DNA BioBank (EADB), Zihuai He, Summer S. Han, Valerio Napolioni, Michael D. Greicius

## Abstract

**Background:** Genetic variants within the *APOE* locus may modulate Alzheimer’s disease (AD) risk independently or in conjunction with *APOE*\*2/3/4 genotypes. Identifying such variants and mechanisms would importantly advance our understanding of *APOE* pathophysiology and provide critical guidance for AD therapies aimed at *APOE*. The *APOE* locus however remains relatively poorly understood in AD, owing to multiple challenges that include its complex linkage structure and uncertainty in *APOE*\*2/3/4 genotype quality. Here, we present a novel *APOE*\*2/3/4 filtering approach and showcase its relevance on AD risk association analyses for the rs439401 variant, which is located 1,801 base pairs downstream of *APOE* and has been associated with a potential regulatory effect on *APOE*.

**Methods:** We used thirty-two AD-related cohorts, with genetic data from various high-density single-nucleotide polymorphism microarrays, whole-genome sequencing, and whole-exome sequencing. Study participants were filtered to be ages 60 and older, non-Hispanic, of European ancestry, and diagnosed as cognitively normal or AD (n=65,701). Primary analyses investigated AD risk in *APOE*\*4/4 carriers. Additional supporting analyses were performed in *APOE*\*3/4 and 3/3 strata. Outcomes were compared under two different *APOE*\*2/3/4 filtering approaches

**Results:** Using more conventional *APOE*\*2/3/4 filtering criteria (approach 1), we showed that, when in-phase with *APOE*\*4, rs439401 was variably associated with protective effects on AD case-control status. However, when applying a novel filter that increases certainty of the *APOE*\*2/3/4 genotypes by applying more stringent criteria for concordance between the provided *APOE* genotype and imputed *APOE* genotype (approach 2), we observed that all significant effects were lost.

**Conclusions:** We showed that careful consideration of *APOE* genotype and appropriate sample filtering were crucial to robustly interrogate the role of the *APOE* locus on AD risk. Our study presents a novel *APOE* filtering approach and provides important guidelines for research into the *APOE* locus, as well as for elucidating genetic interaction effects with *APOE*\*2/3/4.

## Introduction

*APOLIPOPROTEIN E*\*4 (*APOE*\*4) is the strongest genetic risk factor for late-onset Alzheimer’s disease (AD)^1^. In subjects of European ancestry, one copy of *APOE*\*4 increases the risk of a clinical diagnosis of AD by about 3-fold and two copies increases risk by about 12-fold^2,3^. *APOE*\*2 on the other hand decreases risk of AD by about half^3^, while *APOE*\*3 is the reference allele. Beyond the two common missense variants that compose *APOE*\*2/3/4 (rs429358 and rs7412), there may be other coding variants on *APOE* or non-coding regulatory variants in the *APOE* locus that further impact AD risk, either independently or in conjunction with *APOE*\*2/3/4^4–15^. This pertains, by example, to a crucial question in the field: why do some *APOE*\*4 carriers remain asymptomatic even into advanced old age? One possibility is that there may be genetic variants in the *APOE* locus that affect APOE*4 availability and in turn mitigate *APOE*\*4-related risk for AD. Identifying such variants would importantly advance our understanding of *APOE*\*4 pathophysiology and provide critical guidance for AD therapies aimed at *APOE*\*4^16,17^.

Despite its therapeutic promise and three active decades of research, the *APOE* locus remains relatively poorly understood in AD. While there are multiple reasons contributing to this, one prominent one is that the *APOE* locus harbors multiple nearby genes and shows a complex linkage disequilibrium (LD) structure with *APOE*\*2/3/4, making it difficult to identify causal variants and interaction effects^18,19^. Other important reasons are that relevant risk variants may be rare, thus requiring large sample sizes, and, that the quality of the *APOE*\*2/3/4 genotype can bear heavily on correctly identifying interaction effects and causal haplotypes. The latter may be of particular relevance given the plethora of available protein-based (e.g. two-dimensional gel electrophoresis and MALDI-TOF mass spectrometry) and DNA- based methods (e.g. TaqMan assays, high-resolution melting analysis, PCR sequencing, etc.) for *APOE*\*2/3/4 genotyping^20–24^. Importantly, these methods have variable quality and limitations related to the haplotypic nature of *APOE*\*2/3/4. For instance, protein-based assays may suffer from biases in detecting different *APOE* isoforms, while DNA-based assays can be affected by rare variants in the genomic region near *APOE*\*2/3/4 (cf. Huang et al. 2021 for a detailed review)^25^. In turn, cohorts that are commonly included in genetic association studies of AD have used variable *APOE* genotyping methods^26–30^, which has thus led to variable *APOE*\*2/3/4 genotype quality across cohorts used in meta-analyses. The approach used to quality control the *APOE*\*2/3/4 genotype is therefore critical to ensure robust association analyses.

In this study, we present analysis approaches and related findings to guide future research in the *APOE* locus. Specifically, we show findings for a large-scale analysis of rs439401 and its association with AD risk. This variant, located 1801 base pairs downstream of *APOE*, was recently identified as a brain *APOE* splice quantitative trait locus (sQTL) in GTEx^31,32^, spurring our interest to investigate it. We hypothesized it may affect *APOE*\*4 related risk for AD and observed that it is most often seen on the same chromosome copy as *APOE*\*3 (i.e. is in-phase with *APOE*3*), but in rare instances was seen together with *APOE*\*4. We thus stratified analyses according to *APOE*\*3 and *APOE*\*4 genotypes to evaluate whether effects depended on the variant being in-phase with *APOE*\*4. We use analyses on this variant to illustrate how critical it is to have accurate *APOE*\*2/3/4 genotype data. Based on initial analyses using a conventional *APOE* filtering approach and a subsequent robustness assessment, we designed and present a novel *APOE* filtering approach that we believe will be highly relevant to help guide further reproducible research in this area.

## Methods

### Ascertainment of Genotype and Phenotype Data

Genotype data for subjects with AD-related clinical outcome measures were available from thirty-two cohorts, incorporating three sequencing projects^33–56^. Across cohorts, genotyping was performed using various high-density single-nucleotide polymorphism (SNP) microarrays, whole-exome sequencing (WES), and whole-genome sequencing (WGS) (**Table S1**). The discovery samples comprised publicly available case-control (majority), family-based, population-based, and longitudinal cohorts. Independent replication samples, genotyped on SNP microarrays, were available from three large cohorts: the Rotterdam study, a population-based prospective study, the European Alzheimer Disease Initiative (EADI), roughly two-thirds of which is from a prospective population-based study and one third from case-control samples, and the European Alzheimer DNA BioBank (EADB), which collated AD case-control samples from 15 European countries. Ascertainment of genotype/phenotype data for each cohort/project are described in detail elsewhere^33,40–44,46,47,54^. Cross-sample genotype/phenotype harmonization for the discovery samples is summarized in **Supplementary Methods**. Phenotypes from respective cohorts were updated as of March 2021. Data were analyzed between December 2019 and June 2021.

### Genetic Data Quality Control and Processing

Genetic data in the discovery samples underwent standard quality control (QC; Plink v1.9) and ancestry determination (SNPweights v2.1; **Figure S1**)^57^. Only non-Hispanic subjects of European ancestry (representing the vast majority of samples) were selected for processing. Data were restricted to those providing coverage of the rs439401 variant. Principal component analysis of genotyped SNPs provided principal components (PCs) capturing population substructure (PC-AiR, **Figure S2**)^58^. Identity-by-descent (IBD) analyses reliably identified kinship down to 3^rd^ degree relatedness (PC-Relate, **Figure S3**)^58^. Sparse genetic relationship matrices (GRM) were constructed to enable analyses including related individuals^59^. SNP array data were used to perform genotype imputation with regard to the TOPMed imputation reference panel^60,61^. Genetic processing of Rotterdam, EADI, and EADB replication samples is described elsewhere^33,54^. Detailed descriptions of all processing steps are in Supplementary Methods and Table S2.

### Ascertainment of rs439401

The rs439401 variant was originally included in our analyses as it had a cross-cohort genotyping rate >80% in the discovery samples. Genotypes were considered from either the direct call on the SNP array data (i.e. called from probe intensity data), or the call from WGS data. We specifically relied solely on directly genotyped data rather than using imputed data in order to obtain unbiassed results. This choice was additionally motivated reasoning that putative rare haplotypes may not be accurately imputed, particularly when using the commonly younger (non-AD) individuals in imputation reference panels^60,62,63^. Genotype reliability for the variant was verified by cross-correspondence across 3,804 duplicate samples in the discovery and by assessing genotype intensity data on the SNP microarray in EADB.

### Ascertainment of *APOE* genotypes

Throughout, we will refer to *APOE*\*2/3/4 genotypes as *APOE* genotypes. *APOE* genotypes were available from: 1) cohort demographics (i.e. “provided” *APOE)*, which generally had *APOE* genotype status determined through various direct genotyping methods (detailed elsewhere^33,54^), 2) directly from WES/WGS calls, or 3) through imputation of rs429358 (which captures the *APOE*\*4 allele) and rs7412 (which captures the *APOE*\*2 allele). It is relevant to note that rs429358 was never directly available on the SNP microarrays. It is further relevant to note that for the current WES data from ADSP, rs7412 was not available, with only rs429358 being reliably called in most subjects. The WES data could thus be used only to verify subjects with a provided *APOE*\*3/3, 3/4, or 4/4 status (cf. **Supplementary Methods**).

### *APOE* genotype filtering criteria

To our understanding, common criteria across prior studies regarding *APOE* genotypes can be summarized as giving priority to provided *APOE* genotypes when available (as direct genotyping methods are generally considered the gold standard), followed by using *APOE* genotypes derived from rs429358 and rs7412 when directly called on a SNP microarray, followed by inference of *APOE* genotypes through (high quality) imputation of rs429358 and rs7412. There is no clear consensus on whether or how any discrepancies across available *APOE* genotypes for a given subject should be adjudicated. Further, with the recent increasing availability of WGS/WES data in the AD field^42,46,51^, these data can now also be used to verify *APOE* genotypes. When high quality WGS/WES calls are available for rs429358 and rs7412 (i.e. good read depth/quality with a clear reference/alternate allele distribution)^64^, the derived *APOE* genotype may be considered the ground truth. Recent work indeed suggests that a higher *APOE* genotype accuracy can be achieved using next generation sequencing compared to conventional gold standard methods^65^.

#### *APOE* filtering approach 1

Based on the above considerations, we designed criteria to use *APOE* genotypes according to the highest available quality. Specifically, when multiple *APOE* genotypes were available for a given subject, the *APOE* genotype we selected followed the priority of WGS/WES over provided/demographic sources. If *APOE* genotype was only available from provided/demographic sources and was discordant across duplicate samples, then those samples were flagged for exclusion (N=73 out of 1,501 (4.86%) unique subjects). Similarly, the correspondence between *APOE* genotypes derived from WES and WGS across duplicate samples was checked and only showed discordance in five subjects differing for *APOE*\*2/3 and *APOE*\*3/3 genotypes across the WES and WGS data (these subjects were excluded). The final set of samples used for association analyses thus did not display any mismatches in prioritized *APOE* genotypes across duplicates, but in some instances the *APOE* genotype from provided/demographic versus WES/WGS sources differed. *APOE* status as inferred from imputation was entirely ignored, reasoning this was less reliable and that rare haplotypes of potential interest in the *APOE* locus may lead to false imputation of *APOE\**2/3/4 genotype.

### *APOE* filtering approach 2

After further assessment of the initial results, we had concerns about the reliability of *APOE* genotype status in some *APOE*\*4 subjects carrying rs439401 (cf. Results). We therefore expanded the first approach to exclude any subjects who had their prioritized *APOE* genotype determined from provided/demographic *APOE* but were still discordant with their imputed *APOE* genotype (N=632 out of 12,753 (4.96%) in the discovery sample after passing all other filtering steps). Note that imputation scores (R^2^) for rs429358 and rs7412 were never lower than 0.8. Information regarding *APOE* imputation, as well as several correspondence checks across different sources of *APOE* genotypes, are provided in the supplementary and referenced in the results section. An additional check for *APOE* genotype consistency was also performed using newly released sequencing data from the ADSP (NG00067.v5)^66^, processed in May 2021 (cf. **Supplementary Methods** and the Results section).

### Simulations of concordance rates between observed and true *APOE*\*4/4 genotypes

In order to understand potential uncertainty in *APOE*\*4/4 genotypes, we simulated different type I and II error rates for *APOE*\*4/4 status. Type I error rate was defined as the probability, p1, to mis-classify non-*APOE*\*4/4 carriers as *APOE\**4/4. Type II error rate was defined as the probability, p2, to mis-classify *APOE*\*4/4 carriers as non-*APOE\**4/4. We considered a range of true frequencies, f_true_, for *APOE*\*4/4 cases and controls respectively with regard to all cases and controls (that is, all *APOE* strata). This range for f_true_ was centered on observations in the current discovery samples, which should represent a reasonable approximation of expected frequencies in case-control samples. The observed frequency, f_obs_, was then defined as f_true_*(1-p2)+(1-f_true_)*p1. The concordance rate between observed and true *APOE*\*4/4 was finally defined as f_true_*(1-p2)/f_obs_.

### Statistical Analyses

Primary analyses evaluated associations of rs439401 with relative risk for AD in *APOE*\*4/4 carriers using additive genetic models. In additional supporting analyses, associations were evaluated in *APOE*\*3/4 carriers, comparing wild-type (WT) to homozygote (HOM) genotypes, ensuring rs439401 was in-phase with *APOE*\*4. The expectation here was to observe similar but attenuated effects compared to associations in *APOE*\*4/4 carriers. Additional associations were evaluated in *APOE*\*3/4 and 3/3 carriers using additive genetic models, with the expectation of observing little or no effect if associations were conditional on being in-phase with *APOE*\*4. *APOE*\*2/4 carriers were not considered given sample paucity. Analyses were restricted to subjects aged 60 and above, consistent with age cutoffs in prior genetic studies of AD^54^. Replication analyses focused only on evaluating variants in-phase with *APOE*\*4.

Cohorts in the discovery were combined into a single mega-analysis, included related subjects, and outcome measures were adjusted for age, sex, the first five genetic PCs, and the GRM. In replications, models included only unrelated subjects and were not adjusted for the GRM. EADI and Rotterdam further adjusted for the first three genetic PCs, while EADB adjusted for the first 20 genetic PCs and genotyping center. Notably, models in the discovery mega-analyses did not adjust for cohort, reasoning that this may inadvertently diminish power given variable cohort sizes and carrier distributions. This is especially relevant in case of lower frequency variants in the *APOE*\*4/4 stratum, where cohort bins and the number of allele observations become very small.

Associations with AD risk were evaluated under a case-control design using linear mixed-model regression in analyses of related subjects and logistic regression in analyses of unrelated subjects. Additional details for model/inclusion criteria are in **Supplementary Methods**. Association analyses were considered significant below a threshold P-value of 0.05. All analyses were performed in R v3.6.0.

## Results

An overview of the study design and *APOE* filtering approaches is presented in **Figure 1**.

**Figure 1.**
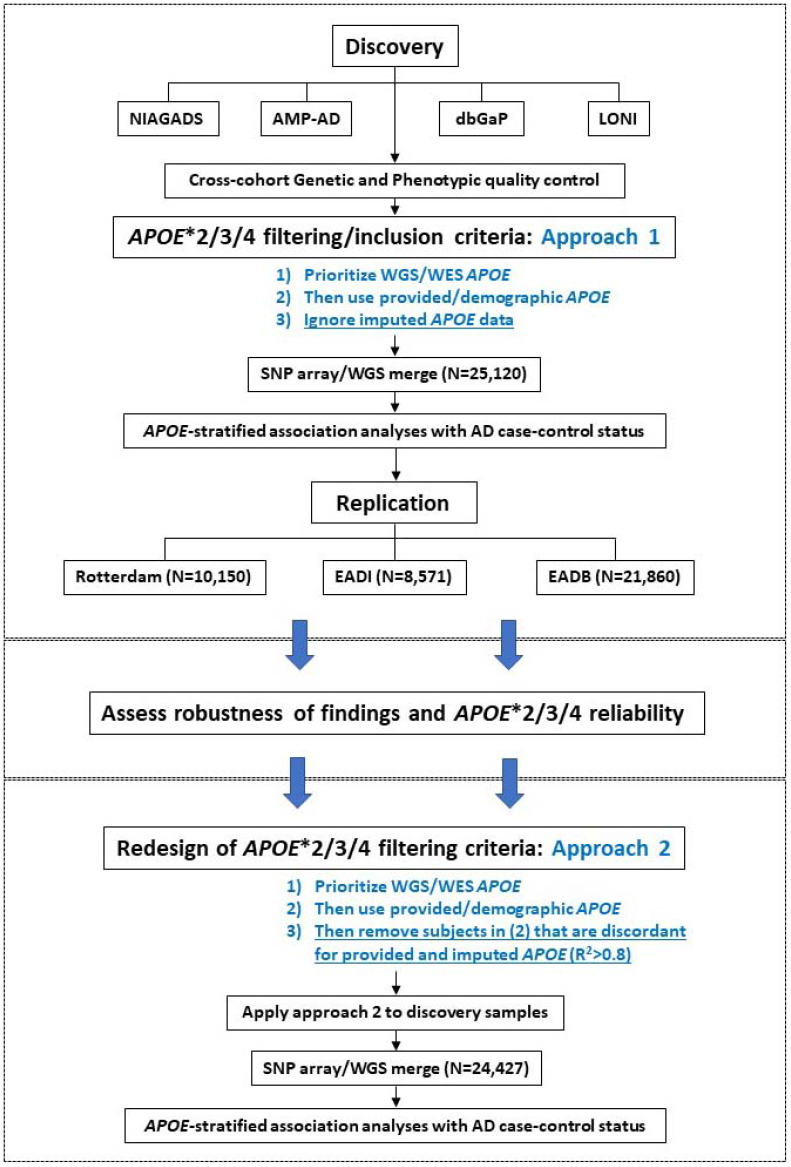
Schematic overview of the study design and two *APOE*\*2/3/4 filtering approaches.

### Participant demographics and rs439401 linkage structure

Across all 142,075 genotyped samples considered in this study (**Table S1**), 65,701 unique participants passed filtering and inclusion criteria. Participant demographics for *APOE*\*4/4 and 3/4 carriers are in **Table 1**, while detailed full sample demographics are in **Table S3-4**. In the discovery, rs439401 displayed high LD (D’>0.9) with *APOE*\*3, but in rarer instances was observed in-phase with *APOE*\*4, thereby deviating from expected LD structure (**Table S5**).

**Table 1.** Sample demographics for association analyses with Alzheimer’s disease case-control status.

| Cohort |  |  |  | APOE *4/4 carriers |  |  | APOE *3/4 carriers |  |  |
| --- | --- | --- | --- | --- | --- | --- | --- | --- | --- |
| Name | Participants after QC (N) | Diagnosis (N) |  | (N (%)) | Female (N (%)) | Age (Mean (SD)) | (N (%)) | Female (N (%)) | Age (Mean (SD)) |
| Discovery | 25120 | CN | 12340 | 238 (1.9 %) | 145 (60.9 %) | 73.7 (7.4) | 238 (1.9 %) | 145 (60.9 %) | 73.7 (7.4) |
|  |  | AD | 12780 | 1652 (12.9 %) | 895 (54.2 %) | 70.1 (6.2) | 1652 (12.9 %) | 895 (54.2 %) | 70.1 (6.2) |
| Replication - Rotterdam | 10150 | CN | 8824 | 150 (1.7 %) | 74 (49.3 %) | 73.8 (8.0) | 150 (1.7 %) | 74 (49.3 %) | 73.8 (8.0) |
|  |  | AD | 1326 | 86 (6.5 %) | 50 (58.1 %) | 78.2 (6.9) | 86 (6.5 %) | 50 (58.1 %) | 78.2 (6.9) |
| Replication - EADI | 8571 | CN | 6502 | 62 (1.0 %) | 45 (72.6 %) | 77.8 (6.4) | 62 (1.0 %) | 45 (72.6 %) | 77.8 (6.4) |
|  |  | AD | 2069 | 205 (9.9 %) | 134 (65.4 %) | 69.4 (5.9) | 205 (9.9 %) | 134 (65.4 %) | 69.4 (5.9) |
| Replication - EADB | 21860 | CN | 12295 | 195 (1.6 %) | 107 (54.9 %) | 70.3 (7.5) | 195 (1.6 %) | 107 (54.9 %) | 70.3 (7.5) |
|  |  | AD | 9565 | 959 (10.0 %) | 551 (57.5 %) | 70.4 (6.8) | 959 (10.0 %) | 551 (57.5 %) | 70.4 (6.8) |

### *APOE* filtering approach 1: Rs439401 shows variable association with Alzheimer’s disease risk

Primary case-control findings in *APOE*\*4/4 carriers in the discovery showed that rs439401 displayed a strong, protective and significant effect on case-control status (**Table 2**). It displayed similar protective effect sizes in EADI and Rotterdam replication samples, but was risk increasing in EADB, and not did not reach significance in any replication sample. When in-phase with *APOE*\*4 in *APOE*\*3/4 (WT-HOM) stratified analyses, rs439401 showed a protective significant effect in the discovery, but variable non-significant results in the replication samples (**Table 2**). In contrast, rs439401 did not associate with AD risk in *APOE*\*3/4 (additive model) or 3/3 stratified analyses in the discovery (**Table S6**).

**Table 2.** Results from *APOE* filtering approach 1: Association findings for rs439401, when in-phase with *APOE* *4, with Alzheimer’s disease case-control status.

| Group/model | Genotype distributions |  |  | AD Case-Control regression |  |
| --- | --- | --- | --- | --- | --- |
|  | CN, carrier<br>No. / Total No. (%) | AD, carrier<br>No. / Total No. (%) | CN - AD, MAF (%) | OR (95% CI) | P-value |
| **rs439401 - T allele tested** |  |  |  |  |  |
| ***APOE* \*4/4 - additive model** |  |  |  |  |  |
| Discovery | 14 / 237 (5.91 %) | 19 / 1652 (1.15 %) | 3.59 % - 0.64 % | 0.10 (0.04, 0.24) | **1.64E-07** |
| Rotterdam | 5 / 150 (3.33 %) | 2 / 86 (2.33 %) | 2.33 % - 1.16 % | 0.43 (0.10, 1.80) | 0.25 |
| EADI | 1 / 62 (1.61 %) | 2 / 205 (0.98 %) | 0.80 % - 0.49 % | 0.37 (0.01, 12.7) | 0.58 |
| EADB | 2 / 195 (1.03 %) | 21 / 956 (2.20 %) | 0.52 % - 1.20 % | 1.61 (0.39, 6.77) | 0.51 |
| ***APOE* \*3/4 - WT vs HOM** |  |  |  |  |  |
| Discovery | 19 / 1401 (1.36 %) | 14 / 2974 (0.47 %) | - | 0.55 (0.38, 0.80) | **1.58E-03** |
| Rotterdam | 8 / 993 (0.81 %) | 3 / 220 (1.36 %) | - | 1.21 (0.31, 4.72) | 0.78 |
| EADI | 4 / 593 (0.67 %) | 5 / 420 (1.19 %) | - | 1.22 (0.60, 2.49) | 0.58 |
| EADB | 12 / 1343 (0.89 %) | 21 / 2070 (1.01 %) | - | 0.80 (0.55, 1.17) | 0.25 |

Abbreviations: CN, cognitively normal; AD, Alzheimer's disease; OR, odds ratio; CI, confidence interval.

Because of the use of a mega-analysis design that does not adjust for cohort, there may still be concern for potential cohort biases. Therefore, as a sensitivity analysis, we re-evaluated the case-control discovery findings, now adjusting for cohort or cohort/array/center (**Figure S5**). These analyses indicated diminished significances, but effect sizes remained comparable and rs439401 remained strongly significant in *APOE*\*4/4 carriers.

### Robustness assessment: Limitations to *APOE* filtering approach 1

After the initial analyses, we assessed the robustness of the primary discovery findings. This appeared particularly relevant considering the very low frequency of rs439401 carriers in *APOE*\*4/4 controls in EADB versus other cohorts, suggesting potential biases in the controls across the cohorts.

Concordance rate of rs439401 from duplicate samples across microarrays and WGS (99.97%) supported genotype reliability (**Table S7**). Similarly, the variant appeared confidently called from the EADB microarray intensity data (**Figure S4**). Overall, we concluded there were no specific genotyping issues for rs439401.

Another important consideration is that some error rate is expected for the different direct *APOE* genotyping methods used across cohorts. Overall, the reliability of the *APOE\**4 genotype may thus be of concern especially when considering the rare *APOE*\*4-rs439401 haplotype. After assessing all *APOE*\*4/4-rs439401 carriers, it was apparent that one cohort, MIRAGE, contributed a large amount of *APOE*\*4/4-rs439401 controls for which *APOE* status was available only from provided/demographic sources (**Figure 2A, Table S8**). We then assessed the concordance rate between provided and imputed *APOE* genotypes across all respective cohorts and observed that MIRAGE displayed the lowest concordance rate of all cohorts included in the discovery analyses (**Figure 2B**), despite comparably high imputation scores for rs429358 and rs7412 to other cohorts (**Table S9**). Overall, this supported concern for the *APOE*\*4/4-rs439401 controls from MIRAGE.

**Figure 2.**
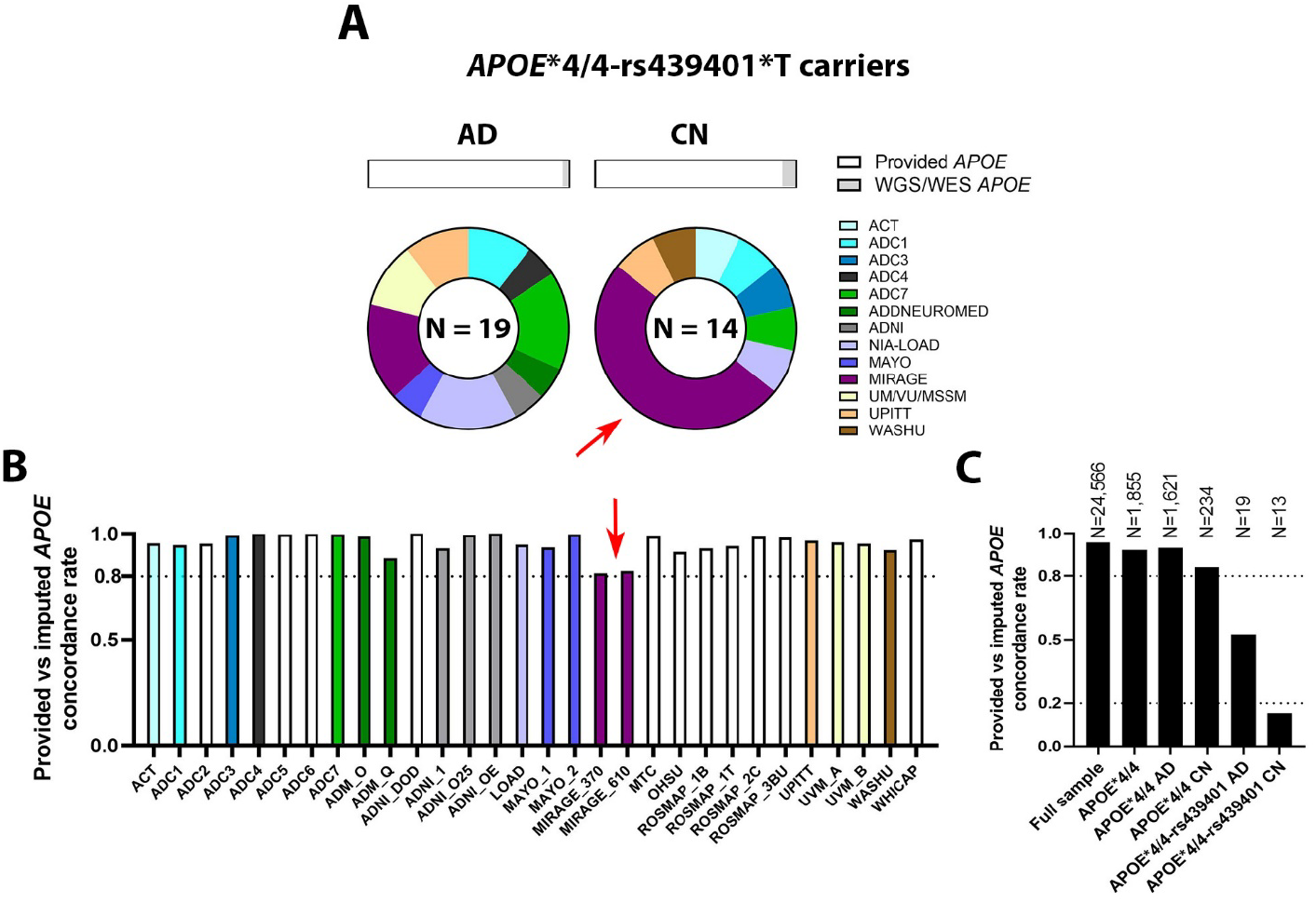
Limitations in *APOE* filtering approach 1 are reflected in discordance between imputed and provided *APOE* genotypes, particularly in *APOE*\*4/4 carriers. **A)** *APOE***\***4/4-rs439401 carrier cohort distributions. Top section shows the distribution of prioritized *APOE* genotype source in approach 1, indicating that *APOE*\*4/4 carriers of rs439401 had very few WGS/WES-verified *APOE*\*4/4 data. Bottom section shows pie charts for carrier distributions across cohorts (additional data in **Table S8**). Red arrow indicates that a large fraction of control rs439401 carriers was contributed by MIRAGE. **B)** Concordance rates between provided and imputed *APOE* per cohort (additional data in **Table S9**). Red arrow indicates that MIRAGE had the lowest concordance rate, suggesting potential limitations with its provided *APOE* data that could explain observations in (A). **C)** Concordance rates between provided and imputed *APOE* for the discovery sample, considering multiple strata (additional data in **Table S10**). *APOE*\*4/4 strata considered provided *APOE*\*4/4 genotypes after applying *APOE* filtering approach 1. Note decreased concordance in *APOE*\*4/4 controls compared to cases. Note strongly decreased concordance for rs439401 carriers, specifically controls. Simulations confirmed that *APOE*\*4/4 controls are more likely than cases to not actually be *APOE*\*4/4 carriers (cf. **Figure S6-7)**. *Abbreviations: CN, cognitively normal; AD, Alzheimer’s disease; OR*.

Extending on the above considerations, we assessed discordance rates between imputed and provided *APOE* for different strata (**Figure 2C, Table S10**). Importantly, while the discordance rate was only 4.3% in the full sample, it increased to 7.2% in *APOE*\*4/4 cases, further increased to 16.1% in *APOE*\*4/4 controls, and then drastically increased to 47.4% in *APOE*\*4/4-rs439401 carrier cases and 85.7% in *APOE*\*4/4-rs439401 carrier controls. While our *a priori* assumption for approach 1 reasoned that imputed *APOE* may be discordant with provided *APOE* in case of subjects with rare haplotypes (e.g. *APOE*\*4/4-rs439401 carriers), the observation that this discordance was 2-fold higher in controls compared to cases would not be expected. Rather, it more likely indicates that a miscall of the *APOE* genotype was true in at least some of these individuals. To better understand these observations, we performed simulation studies using different type I and type II error rates (0-5%) for *APOE*\*4/4 genotyping and observed that *APOE*\*4/4 controls were more likely than *APOE*\*4/4 cases to not actually be *APOE*\*4/4 carriers (**Figure S6-7**). This was the result of the low frequency of *APOE*\*4/4 controls and the strong case-control imbalance in *APOE*\*4/4 carriers. Overall, this supported concern for the validity of approach 1.

We then used the recently released new ADSP WGS and WES data, which now cover additional subjects that are duplicated on SNP array samples included in our discovery analyses (N=3,644 as determined by identity-by-descent). We assessed the *APOE* genotype calls from the novel WES/WGS data and observed that three *APOE*\*4/4-rs439401 control subjects (not from the MIRAGE cohort) in the prior discovery samples were in fact *APOE*\*3/3 or *APOE*\*3/4 carriers, which was also the imputed *APOE* genotype (**Table S8**). Overall, this again raised concern about the validity of approach 1.

In sum, these additional checks for robustness of the findings suggested problems with *APOE* genotype reliability in subjects with *APOE*\*4-rs439401 haplotypes and *APOE*\*4/4 carriers overall, indicating a limitation to the first (conventional) *APOE* filtering approach. In a final check, we observed that despite good concordance between provided and WGS *APOE* (99.1%), imputed and WGS *APOE* was more concordant (97.2%) than imputed and provided *APOE* (95.7%), indicating that at least in some subjects imputed *APOE* is likely more correct than provided *APOE* (**Table S10**).

### *APOE* filtering approach 2: Rs439401 shows no association with Alzheimer’s disease risk

In light of the identified *APOE* reliability limitations, we extended approach 1 to filter out any subjects that did not have WGS/WES *APOE* and at the same time were discordant for provided and (high-quality) imputed *APOE*. We also filtered out any discordant *APOE* calls with regard to the new ADSP WES/WGS data since this information was available (in case of *APOE*\*4-rs439401 carriers, this overlapped with samples where provided and imputed *APOE* were discordant). We then applied this to the discovery samples and reran analyses. Exclusion of subjects with discordant *APOE* status with the newly released ASDP WES/WGS data removed 61 (out of 12,367) subjects from the SNP-array samples. Further applying the new *APOE* filter excluded 632 (out of 12,753 considered) subjects from the discovery SNP-array samples. *APOE*\*4-rs439401 carrier frequencies dropped substantially, particularly in controls, and became more consistent with those observed in the haplotype reference consortium (**Figure 3A**). Case-control association analyses now indicated no effects for *APOE*\*4-rs439401 carriers (**Table 3 & Figure 3B-C**). In sum, approach 2 produced results that were more realistic in terms of expected linkage structure and more consistent with the lack of significant replication findings.

**Table 3.** Results from *APOE* filtering approach 1 versus 2 in the discovery: Association findings for rs439401, when in-phase with *APOE* *4, with Alzheimer’s disease case-control status.

| Group/model | Genotype distributions |  |  | AD Case-Control regression |  |
| --- | --- | --- | --- | --- | --- |
|  | CN, carrier<br>No. / Total No. (%) | AD, carrier<br>No. / Total No. (%) | CN - AD, MAF (%) | OR (95% CI) | P-value |
| **rs439401 - T allele tested** |  |  |  |  |  |
| ***APOE* \*4/4 - additive model** |  |  |  |  |  |
| Discovery - approach 1 | 14 / 237 (5.91 %) | 19 / 1652 (1.15 %) | 3.59 % - 0.64 % | 0.10 (0.04, 0.24) | **1.64E-07** |
| Discovery - approach 2 | 2 / 203 (0.99 %) | 10 / 1577 (0.63 %) | 0.49 % - 0.32 % | 0.47 (0.07, 3.21) | 0.44 |
| ***APOE* \*3/4 - WT vs HOM** |  |  |  |  |  |
| Discovery - approach 1 | 19 / 1401 (1.36 %) | 14 / 2974 (0.47 %) | - | 0.55 (0.38, 0.80) | **1.58E-03** |
| Discovery - approach 2 | 4 / 1339 (0.29 %) | 9 / 2914 (0.31 %) | - | 1.09 (0.61, 1.94) | 0.78 |

*Abbreviations: CN, cognitively normal; AD, Alzheimer's disease; OR, odds ratio; CI, confidence interval.*

**Figure 3.**
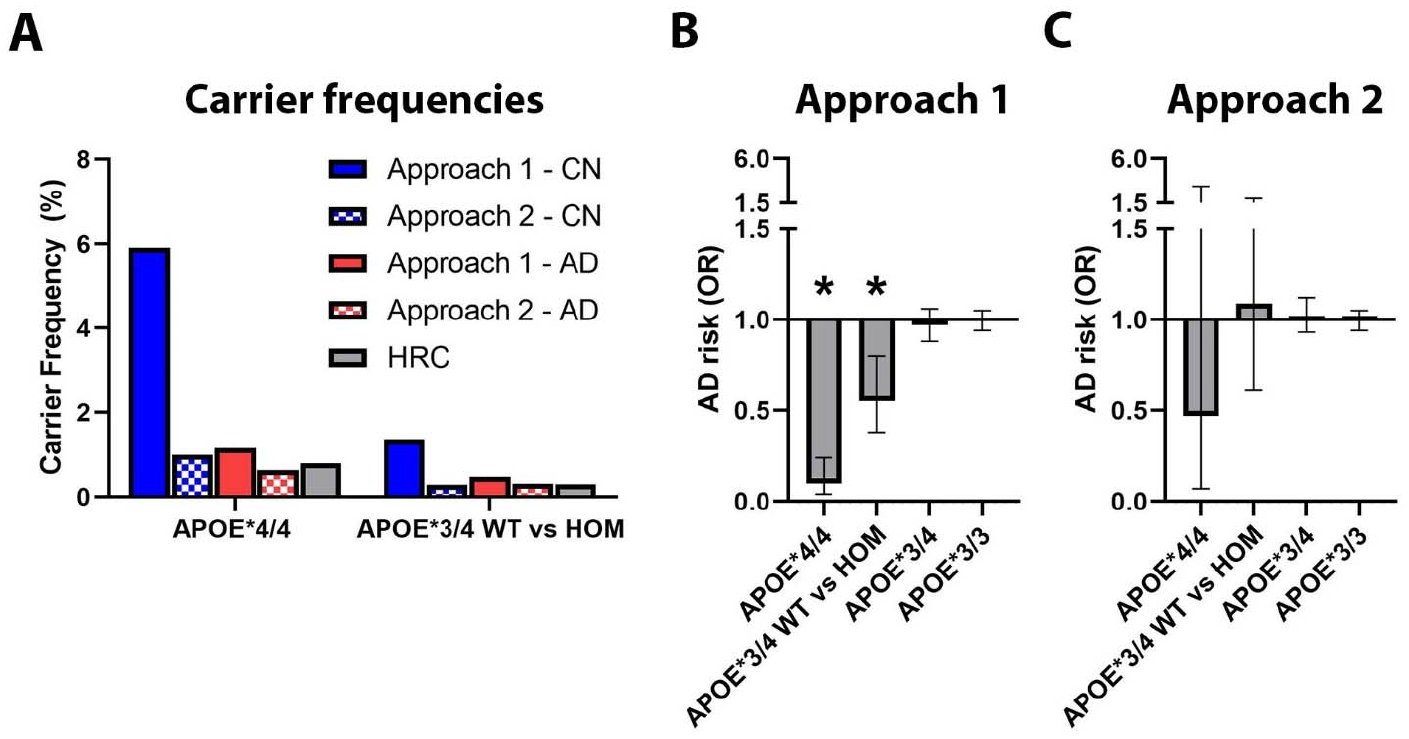
Overview of rs439401 frequencies and case-control association findings, comparing *APOE* filtering approach 1 to approach 2. **A)** Carrier frequencies across both approaches for *APOE*\*4/4 and *APOE*\*3/4 WT vs HOM groups, as well as in the Haplotype reference consortium v1.1 (HRC). Note decreased frequencies for rs439401 in approach 2 that appear concordant with the HRC. **B-C)** Overview of association findings for all evaluated strata, comparing **B)** approach 1 to **C)** approach 2. Significant effects are denoted by an asterisk (*). Error bars show 95% confidence intervals. Note loss of significant effects in approach 2.

## Discussion

Our results demonstrate that the filtering criteria for *APOE*\*2/3/4 genotypes can heavily impact association finding for variants that exert their effect in conjunction with *APOE*\*2/3/4. Specifically, we used the *APOE* sQTL variant rs439401 to illustrate this point. Using more conventional filtering criteria regarding *APOE* genotypes (approach 1), we showed that, when in-phase with *APOE*\*4, rs439401 was variably associated with protective effects on AD case-control status. However, when assessing the reliability of *APOE*\*2/3/4 genotypes with more scrutiny and applying a novel filter to increase certainty of the *APOE* genotypes (approach 2), we observed that all significant effects were lost. The findings and methodology presented here are thus of high relevance to guide future research into the *APOE* locus.

The rs439401 variant considered in the current study has previously been investigated with regard to AD risk in different contexts and using variable strategies and study designs^8–11,13^. Our analyses however considered a substantially larger sample size, essentially incorporating most European ancestry AD cohorts included in prior studies, specifically focused on evaluating effects stratified to respective *APOE* genotypes, and tested only directly genotyped variants. Further, up-to-date genotype and phenotype data for a large set of AD cohorts was jointly harmonized to compose a parsimonious discovery sample. Non-European ancestries were not investigated here owing to the paucity of publicly available data. When compared to similar prior studies^6,13–15^, our discovery group was larger and we incorporated three large replication cohorts. Further, through the implementation of linear mixed modeling and cross-sample harmonization, we were able to increase the power and specificity for variant discovery, while additionally verifying genotype reliability across nearly 4,000 duplicate samples. In sum, our analyses should provide a robust assessment of the presented *APOE* filtering approaches and rs439401’s association with AD risk.

A recent study, using samples largely overlapping with the current discovery (but smaller in size) and an *APOE* filtering approach similar to our approach 1, evaluated the association of variants on the larger *APOE* locus with AD risk in *APOE*\*4/4 carriers and did not identify the strong association of rs439401 that we observed in approach 1^13^. Beyond differences in sample size and harmonization, the latter study adjusted models by study/cohort and made use of imputed genotypes. We specifically decided in primary analyses not to adjust for cohort, as we reasoned that this may inadvertently diminish power given variable cohort sizes and carrier distributions, especially in *APOE*\*4/4 carriers. We further reasoned that through our extensive phenotype/genotype harmonization and the use of a mixed model mega-analysis design, which may capture some latent cohort effects, there was less concern for potential cohort bias. Additionally, given the complex LD structure of the *APOE* locus, we were concerned about the reliability of imputation and focused only on directly called genotypes. A similar limitation regarding imputation was recognized by the authors of the prior study^13^. These differences likely explain why rs439401 was not observed in their study. Regardless of our considerations and of cohort adjustment, we determined that the *APOE* filtering criteria were the most relevant factor for variable rs439401 association findings.

One important insight from our study was that subjects, particularly controls, with a provided *APOE*\*4/4 genotype had a higher probability of discordance between their imputed and provided genotype than did subjects in the full sample. Such biases are, however, not limited to *APOE*\*4/4 carriers. The six *APOE* genotypes (*2/2, 2/3, 3/3, 2/4, 3/4, 4/4) show large differences in numbers of carriers and case-control ratios, owing to the allele frequencies of rs429358/rs7412 and their effect on AD risk. As a result, the different *APOE* genotypes will be expected to have different concordance rates between true and observed *APOE* genotypes. We observed varying concordance between imputed and provided *APOE* across the six *APOE* genotypes, with particularly lower concordance rates in *APOE*\*2 carriers (**Figure S8**). Just as the *APOE\**4/4 provided genotype was most likely to be incorrect here in controls (a phenotype for which *APOE\**4/4 is a particularly rare genotype), the *APOE\**2/2 genotype is more likely to be incorrect in cases (a phenotype for which *APOE\**2/2 is a particularly rare genotype). The proposed *APOE* genotype filter will therefore also be specifically relevant for studies focusing on *APOE*\*2.

Our study highlights several important considerations for further work on the *APOE* locus. Most notably, we illustrate how *APOE* genotype filtering criteria can strongly impact association findings for variants in the *APOE* locus, especially when studying haplotypes or interaction effects with *APOE*\*2/3/4. The same will hold true when considering non-local variants in, for instance, a genome-wide association study of AD in *APOE*\*4/4 subjects, or when aiming to disentangle genetic interaction effects with *APOE*\*2/3/4. Based on our observations, we suggest that future studies consider implementing the methodology that we proposed in approach 2 and subject their assessment of *APOE* genotypes to extensive scrutiny. The limitations observed for *APOE*\*2/3/4 genotype reliability also emphasize that next generation sequencing data will be crucial to interrogate the *APOE* locus with higher confidence and to ensure that putative rare haplotypes are not missed because of the need for sample filtering in SNP array data. Lastly, in order to have higher confidence in local haplotypes, long read sequencing approaches will additionally be crucial to help disentangle the local haplotype structure on *APOE* with regard to AD.

## Conclusion

We showed that careful consideration of *APOE* genotype and appropriate sample filtering was crucial to robustly interrogate the role of the *APOE* locus on AD risk. Our study presents a novel *APOE* filtering approach and provides important guidelines for research in this area, as well as for elucidating genetic interaction effects with *APOE*\*2/3/4.

## Data Availability

All data used in the discovery analyses are available upon application to:
- dbGaP (https://www.ncbi.nlm.nih.gov/gap/)
- NIAGADS (https://www.ncbi.nlm.nih.gov/gap/)
- LONI (https://ida.loni.usc.edu/)
- Synapse (https://www.synapse.org/)
- Rush (https://www.radc.rush.edu/)
- NACC (https://naccdata.org/)
The specific data repository and identifier for each cohort is indicated in Table S1 of the supplement. Because of ethical and legal restrictions, the Rotterdam, EADI, and EADB cohort data are available only upon request. Interested authors may respectively contact Dr. Ikram, at, and Dr. Lambert, at.

https://www.ncbi.nlm.nih.gov/gap/

https://ida.loni.usc.edu/

https://www.synapse.org/

https://www.radc.rush.edu/

https://naccdata.org/

## Ethics approval and consent to participate

Participants or their caregivers provided written informed consents in the original studies. The current study protocol was granted an exemption by the Stanford Institutional Review Board because the analyses were carried out on “de-identified, off-the-shelf” data.

## Consent for publication

Not applicable.

## Availability of data and materials

All data used in the discovery analyses are available upon application to:

- dbGaP (https://www.ncbi.nlm.nih.gov/gap/)
- NIAGADS (https://www.niagads.org/)
- LONI (https://ida.loni.usc.edu/)
- Synapse (https://www.synapse.org/)
- Rush (https://www.radc.rush.edu/)
- NACC (https://naccdata.org/)

The specific data repository and identifier for each cohort is indicated in Table S1 of the supplement. Because of ethical and legal restrictions, the Rotterdam, EADI, and EADB cohort data are available only upon request. Interested authors may respectively contact Dr. Ikram, at, and Dr. Lambert, at.

## Authors’ contributions

M.E.B. performed data processing, performed data analyses, designed analyses, designed study, wrote paper, and obtained funding. S.J.E. performed data processing and performed data analyses. Y.L. performed data processing. S.A. and V.D. performed data processing and performed data analyses. M.A.I and J.-C.L. contributed data and supervised analyses for the Rotterdam and EADI/EADB cohorts respectively. A.R, A.C.T, G.R, I.E.J, I.d.R, K.P, K.S., M.I., M.H. N.A., O.A., P.S.J, P.K, P.A., R.S., R.F-S, W.M.v.d.F contributed data for the EADB cohort. Z.H. designed analyses and supervised analyses. V.N. performed data processing and supervised work. S.S.H. designed study, designed analyses, and supervised analyses. M.D.G designed study, designed analyses, supervised analyses, supervised work, wrote paper, and obtained funding.

## Acknowledgements

NCRAD. Biological samples used in this study were stored at study investigators’ institutions and at the National Cell Repository for Alzheimer’s Disease (NCRAD) at Indiana University, which receives government support under a cooperative agreement grant (U24 AG21886) awarded by the National Institute on Aging (NIA). We thank contributors who collected samples used in this study, as well as patients and their families, whose help and participation made this work possible.

Phenotypic data were provided by principal investigators, the NIA funded Alzheimer’s Disease Centers (ADCs), the National Alzheimer’s Coordinating Center (NACC, U01AG016976), and the National Institute on Aging Genetics of Alzheimer’s Disease Data Storage Site (NIAGADS, U24AG041689) at the University of Pennsylvania, funded by NIA. Contributors to the Genetic Analysis Data included Study Investigators on projects that were individually funded by NIA, and other NIH institutes, and by private U.S. organizations, or foreign governmental or nongovernmental organizations.

NIAGADS/ADGC. Data for this study were prepared, archived, and distributed by the National Institute on Aging Alzheimer’s Disease Data Storage Site (NIAGADS) at the University of Pennsylvania (U24- AG041689-01); Alzheimer’s Disease Genetics Consortium (ADGC), U01 AG032984, RC2 AG036528; NACC, U01 AG016976; NIA-LOAD (Columbia University), U24 AG026395, U24 AG026390, R01AG041797; Banner Sun Health Research Institute P30 AG019610; Boston University, P30 AG013846, U01 AG10483, R01 CA129769, R01 MH080295, R01 AG017173, R01 AG025259, R01 AG048927, R01AG33193, R01 AG009029; Columbia University, P50 AG008702, R37 AG015473, R01 AG037212, R01 AG028786; Duke University, P30 AG028377, AG05128; Emory University, AG025688; Group Health Research Institute, UO1 AG006781, UO1 HG004610, UO1 HG006375, U01 HG008657; Indiana University, P30 AG10133, R01 AG009956, RC2 AG036650; Johns Hopkins University, P50 AG005146, R01 AG020688; Massachusetts General Hospital, P50 AG005134; Mayo Clinic, P50 AG016574, R01 AG032990, KL2 RR024151; Mount Sinai School of Medicine, P50 AG005138, P01 AG002219; New York University, P30 AG08051, UL1 RR029893, 5R01AG012101, 5R01AG022374, 5R01AG013616, 1RC2AG036502, 1R01AG035137; North Carolina A&T University, P20 MD000546, R01 AG28786-01A1; Northwestern University, P30 AG013854; Oregon Health & Science University, P30 AG008017, R01 AG026916; Rush University, P30 AG010161, R01 AG019085, R01 AG15819, R01 AG17917, R01 AG030146, R01 AG01101, RC2 AG036650, R01 AG22018; TGEN, R01 NS059873; University of Alabama at Birmingham, P50 AG016582, UL1RR02777; University of Arizona, R01 AG031581; University of California, Davis, P30 AG010129; University of California, Irvine, P50 AG016573, P50 AG016575, P50 AG016576, P50 AG016577; University of California, Los Angeles, P50 AG016570; University of California, San Diego, P50 AG005131; University of California, San Francisco, P50 AG023501, P01 AG019724; University of Kentucky, P30 AG028383, AG05144; University of Michigan, P30 AG053760 and AG063760; University of Pennsylvania, P30 AG010124; University of Pittsburgh, P50 AG005133, AG030653, AG041718, AG07562, AG02365; University of Southern California, P50 AG005142; University of Texas Southwestern, P30 AG012300; University of Miami, R01 AG027944, AG010491, AG027944, AG021547, AG019757; University of Washington, P50 AG005136, R01 AG042437; University of Wisconsin, P50 AG033514; Vanderbilt University, R01 AG019085; and Washington University, P50 AG005681, P01 AG03991, P01 AG026276. The Kathleen Price Bryan Brain Bank at Duke University Medical Center is funded by NINDS grant # NS39764, NIMH MH60451 and by Glaxo Smith Kline. Genotyping of the TGEN2 cohort was supported by Kronos Science. The TGen series was also funded by NIA grant AG041232, The Banner Alzheimer’s Foundation, The Johnnie B. Byrd Sr. Alzheimer’s Institute, the Medical Research Council, and the state of Arizona and also includes samples from the following sites: Newcastle Brain Tissue Resource (funding via the Medical Research Council, local NHS trusts and Newcastle University), MRC London Brain Bank for Neurodegenerative Diseases (funding via the Medical Research Council), South West Dementia Brain Bank (funding via numerous sources including the Higher Education Funding Council for England (HEFCE), Alzheimer’s Research Trust (ART), BRACE as well as North Bristol NHS Trust Research and Innovation 58 Department and DeNDRoN), The Netherlands Brain Bank (funding via numerous sources including Stichting MS Research, Brain Net Europe, Hersenstichting Nederland Breinbrekend Werk, International Parkinson Fonds, Internationale Stiching Alzheimer Onderzoek), Institut de Neuropatologia, Servei Anatomia Patologica, Universitat de Barcelona.

NACC. The NACC database is funded by NIA/NIH Grant U01 AG016976. NACC data are contributed by the NIA-funded ADCs: P30 AG019610 (PI Eric Reiman, MD), P30 AG013846 (PI Neil Kowall, MD), P30 AG062428-01 (PI James Leverenz, MD) P50 AG008702 (PI Scott Small, MD), P50 AG025688 (PI Allan Levey, MD, PhD), P50 AG047266 (PI Todd Golde, MD, PhD), P30 AG010133 (PI Andrew Saykin, PsyD), P50 AG005146 (PI Marilyn Albert, PhD), P30 AG062421-01 (PI Bradley Hyman, MD, PhD), P30 AG062422- 01 (PI Ronald Petersen, MD, PhD), P50 AG005138 (PI Mary Sano, PhD), P30 AG008051 (PI Thomas Wisniewski, MD), P30 AG013854 (PI Robert Vassar, PhD), P30 AG008017 (PI Jeffrey Kaye, MD), P30 AG010161 (PI David Bennett, MD), P50 AG047366 (PI Victor Henderson, MD, MS), P30 AG010129 (PI Charles DeCarli, MD), P50 AG016573 (PI Frank LaFerla, PhD), P30 AG062429-01(PI James Brewer, MD, PhD), P50 AG023501 (PI Bruce Miller, MD), P30 AG035982 (PI Russell Swerdlow, MD), P30 AG028383 (PI Linda Van Eldik, PhD), P30 AG053760 (PI Henry Paulson, MD, PhD), P30 AG010124 (PI John Trojanowski, MD, PhD), P50 AG005133 (PI Oscar Lopez, MD), P50 AG005142 (PI Helena Chui, MD), P30 AG012300 (PI Roger Rosenberg, MD), P30 AG049638 (PI Suzanne Craft, PhD), P50 AG005136 (PI Thomas Grabowski, MD), P30 AG062715-01 (PI Sanjay Asthana, MD, FRCP), P50 AG005681 (PI John Morris, MD), P50 AG047270 (PI Stephen Strittmatter, MD, PhD).

GenADA. The genotypic and associated phenotypic data used in the study “Multi-Site Collaborative Study for Genotype-Phenotype Associations in Alzheimer’s Disease (GenADA)” were provided by the GlaxoSmithKline, R&D Limited.

ROSMAP. ROSMAP study data were provided by the Rush Alzheimer’s Disease Center, Rush University Medical Center, Chicago. Data collection was supported through funding by NIA grants P30AG10161, R01AG15819, R01AG17917, R01AG30146, R01AG36836, U01AG32984, U01AG46152, the Illinois Department of Public Health, and the Translational Genomics Research Institute.

AddNeuroMed. The AddNeuroMed data are from a public-private partnership supported by EFPIA companies and SMEs as part of InnoMed (Innovative Medicines in Europe), an Integrated Project funded by the European Union of the Sixth Framework program priority FP6-2004-LIFESCIHEALTH-5. Clinical leads responsible for data collection are Iwona Kłoszewska (Lodz), Simon Lovestone (London), Patrizia Mecocci (Perugia), Hilkka Soininen (Kuopio), Magda Tsolaki (Thessaloniki), and Bruno Vellas (Toulouse), imaging leads are Andy Simmons (London), Lars-Olad Wahlund (Stockholm) and Christian Spenger (Zurich) and bioinformatics leads are Richard Dobson (London) and Stephen Newhouse (London).

ADNI. Data collection and sharing for this project was funded by the Alzheimer’s Disease Neuroimaging Initiative (ADNI) (National Institutes of Health Grant U01 AG024904) and DOD ADNI (Department of Defense award number W81XWH-12-2-0012). ADNI is funded by the National Institute on Aging, the National Institute of Biomedical Imaging and Bioengineering and through generous contributions from the following: AbbVie. Alzheimer’s Association; Alzheimer’s Drug Discovery Foundation; Araclon Biotech; BioClinica. Inc.; Biogen; Bristol-Myers Squibb Company; CereSpir. Inc.; Cogstate; Eisai Inc.; Elan Pharmaceuticals. Inc.; Eli Lilly and Company; EuroImmun; F. Hoffmann-La Roche Ltd and its affiliated company Genentech. Inc.; Fujirebio; GE HealtControlsare; IXICO Ltd.; Janssen Alzheimer Immunotherapy Research & Development. LLC.; Johnson & Johnson Pharmaceutical Research & Development LLC.; Lumosity; Lundbeck; Merck & Co. Inc.; Meso Scale Diagnostics. LLC.; NeuroRx Research; Neurotrack Technologies; Novartis Pharmaceuticals Corporation; Pfizer Inc.; Piramal Imaging; Servier; Takeda Pharmaceutical Company; and Transition Therapeutics. The Canadian Institutes of Health Research is providing funds to support ADNI clinical sites in Canada. Private sector contributions are facilitated by the Foundation for the National Institutes of Health. The grantee organization is the Northern California Institute for Research and Education, and the study is coordinated by the Alzheimer’s Therapeutic Research Institute at the University of Southern California. ADNI data are disseminated by the Laboratory for Neuro Imaging at the University of Southern California.

EADB. We thank the many study participants, researchers, and staff for collecting and contributing to the data, the high-performance computing service at the University of Lille, and the staff at CEA-CNRGH for their help with sample preparation and genotyping, and excellent technical assistance. We thank Antonio Pardinas for his help. This research was conducted using the UK Biobank resource (application number: 61054).

This work was funded by a grant (European Alzheimer DNA BioBank, EADB) from the EU Joint Programme – Neurodegenerative Disease Research (JPND). Inserm UMR1167 is also funded by the Inserm, Institut Pasteur de Lille, Lille Métropole Communauté Urbaine, and the French government’s LABEX DISTALZ program (development of innovative strategies for a transdisciplinary approach to Alzheimer’s disease).

Additional support for EADB cohorts. The work for this manuscript was further supported by the CoSTREAM project (www.costream.eu) and funding from the European Union’s Horizon 2020 research and innovation programme under grant agreement No 667375. This work is also funded by la fondation pour la recherché médicale (FRM) (EQU202003010147) Italian Ministry of Health (Ricerca Corrente); Ministero dell’Istruzione, del l’Università e della Ricerca–MIUR project “Dipartimenti di Eccellenza 2018– 2022” to Department of Neuroscience “Rita Levi Montalcini”, University of Torino (IR), and AIRAlzh Onlus-ANCC-COOP (SB); Partly supported by “Ministero della Salute”, I.R.C.C.S. Research Program, Ricerca Corrente 2018-2020, Linea n. 2 “Meccanismi genetici, predizione e terapie innovative delle malattie complesse” and by the “5 × 1000” voluntary contribution to the Fondazione I.R.C.C.S. Ospedale “Casa Sollievo della Sofferenza”; and RF-2018-12366665, Fondi per la ricerca 2019 (Sandro Sorbi). Copenhagen General Population Study (CGPS): We thank staff and participants of the CGPS for their important contributions. Karolinska Institutet AD cohort: Dr. Graff and co-authors of the Karolinska Institutet AD cohort report grants from Swedish Research Council (VR) 2015-02926, 2018-02754, 2015- 06799, Swedish Alzheimer Foundation, Stockholm County Council ALF and resarch school, Karolinska Institutet StratNeuro, Swedish Demensfonden, and Swedish brain foundation, during the conduct of the study. ADGEN: This work was supported by Academy of Finland (grant numbers 307866); Sigrid Jusélius Foundation; the Strategic Neuroscience Funding of the University of Eastern Finland; EADB project in the JPNDCO-FUND program (grant number 301220). CBAS: Supported by the project no. LQ1605 from the National Program of Sustainability II (MEYS CR), Supported by Ministry of Health of the Czech Republic, grant nr. NV19-04-00270 (All rights reserved), Grant Agency of Charles University Grants No. 693018 and 654217; the Ministry of Health, Czech Republic―conceptual development of research organization, University Hospital Motol, Prague, Czech Republic Grant No. 00064203; the Czech Ministry of Health Project AZV Grant No. 16―27611A; and Institutional Support of Excellence 2. LF UK Grant No. 699012. CNRMAJ-Rouen: This study received fundings from the Centre National de Référence Malades Alzheimer Jeunes (CNRMAJ). The Finnish Geriatric Intervention Study for the Prevention of Cognitive Impairment and Disability (FINGER) data collection was supported by grants from the Academy of Finland, La Carita Foundation, Juho Vainio Foundation, Novo Nordisk Foundation, Finnish Social Insurance Institution, Ministry of Education and Culture Research Grants, Yrjö Jahnsson Foundation, Finnish Cultural Foundation South Osthrobothnia Regional Fund, and EVO/State Research Funding grants of University Hospitals of Kuopio, Oulu and Turku, Seinäjoki Central Hospital and Oulu City Hospital, Alzheimer’s Research & Prevention Foundation USA, AXA Research Fund, Knut and Alice Wallenberg Foundation Sweden, Center for Innovative Medicine (CIMED) at Karolinska Institutet Sweden, and Stiftelsen Stockholms sjukhem Sweden. FINGER cohort genotyping was funded by EADB project in the JPND CO- FUND (grant number 301220). Research at the Belgian EADB site is funded in part by the Alzheimer Research Foundation (SAO-FRA), The Research Foundation Flanders (FWO), and the University of Antwerp Research Fund. FK is supported by a BOF DOCPRO fellowship of the University of Antwerp Research Fund. SNAC-K is financially supported by the Swedish Ministry of Health and Social Affairs, the participating County Councils and Municipalities, and the Swedish Research Council. BDR Bristol: We would like to thank the South West Dementia Brain Bank (SWDBB) for providing brain tissue for this study. The SWDBB is part of the Brains for Dementia Research programme, jointly funded by Alzheimer’s Research UK and Alzheimer’s Society and is supported by BRACE (Bristol Research into Alzheimer’s and Care of the Elderly) and the Medical Research Council. BDR Manchester: We would like to thank the Manchester Brain Bankfor providing brain tissue for this study. The Manchester Brain Bank is part of the Brains for Dementia Research programme, jointly funded by Alzheimer’s Research UK and Alzheimer’s Society. BDR KCL: Human post-mortem tissue was provided by the London Neurodegenerative Diseases Brain Bank which receives funding from the UK Medical Research Council and as part of the Brains for Dementia Research programme, jointly funded by Alzheimer’s Research UK and the Alzheimer’s Society. The CFAS Wales study was funded by the ESRC (RES-060-25-0060) and HEFCW as ‘Maintaining function and well-being in later life: a longitudinal cohort study’, (Principal Investigators: R.T Woods, L.Clare, G.Windle, V. Burholt, J. Philips, C. Brayne, C. McCracken, K. Bennett, F. Matthews). We are grateful to the NISCHR Clinical Research Centre for their assistance in tracing participants and in interviewing and in collecting blood samples, and to general practices in the study areas for their cooperation. MRC: We thank all individuals who participated in this study. Cardiff University was supported by the Alzheimer’s Society (AS; grant RF014/164) and the Medical Research Council (MRC; grants G0801418/1, MR/K013041/1, MR/L023784/1) (R. Sims is an AS Research Fellow). Cardiff University was also supported by the European Joint Programme for Neurodegenerative Disease (JPND; grant MR/L501517/1), Alzheimer’s Research UK (ARUK; grant ARUK-PG2014-1), the Welsh Assembly Government (grant SGR544:CADR), Brain’s for dementia Research and a donation from the Moondance Charitable Foundation. Cardiff University acknowledges the support of the UK Dementia Research Institute, of which J. Williams is an associate director. Cambridge University acknowledges support from the MRC. Patient recruitment for the MRC Prion Unit/UCL Department of Neurodegenerative Disease collection was supported by the UCLH/UCL Biomedical Centre and NIHR Queen Square Dementia Biomedical Research Unit. The University of Southampton acknowledges support from the AS. King’s College London was supported by the NIHR Biomedical Research Centre for Mental Health and the Biomedical Research Unit for Dementia at the South London and Maudsley NHS Foundation Trust and by King’s College London and the MRC. ARUK and the Big Lottery Fund provided support to Nottingham University. Alfredo Ramirez: Part of the work was funded by the JPND EADB grant (German Federal Ministry of Education and Research (BMBF) grant: 01ED1619A). Alfredo Ramirez is also supported by the German Research Foundation (DFG) grants Nr: RA 1971/6-1, RA1971/7-1, and RA 1971/8-1. German Study on Ageing, Cognition and Dementia in Primary Care Patients (AgeCoDe): This study/publication is part of the German Research Network on Dementia (KND), the German Research Network on Degenerative Dementia (KNDD; German Study on Ageing, Cognition and Dementia in Primary Care Patients; AgeCoDe), and the Health Service Research Initiative (Study on Needs, health service use, costs and health-related quality of life in a large sample of oldestold primary care patients (85+; AgeQualiDe)) and was funded by the German Federal Ministry of Education and Research (grants KND: 01GI0102, 01GI0420, 01GI0422, 01GI0423, 01GI0429, 01GI0431, 01GI0433, 01GI0434; grants KNDD: 01GI0710, 01GI0711, 01GI0712, 01GI0713, 01GI0714, 01GI0715, 01GI0716; grants Health Service Research Initiative: 01GY1322A, 01GY1322B, 01GY1322C, 01GY1322D, 01GY1322E, 01GY1322F, 01GY1322G). VITA study: The support of the Ludwig Boltzmann Society and the AFI Germany have supported the VITA study. The former VITA study group should be acknowledged: W. Danielczyk, G. Gatterer, K Jellinger, S Jugwirth, KH Tragl, S Zehetmayer. Vogel Study: This work was financed by a research grant of the ‘‘Vogelstiftung Dr. Eckernkamp’’. HELIAD study: This study was supported by the grants: IIRG-09-133014 from the Alzheimer’s Association, 189 10276/8/9/2011 from the ESPA-EU program Excellence Grant (ARISTEIA) and the ΔY2β/οικ.51657/14.4.2009 of the Ministry for Health and Social Solidarity (Greece). Biobank Department of Psychiatry, UMG: Prof. Jens Wiltfang is supported by an Ilídio Pinho professorship and iBiMED (UID/BIM/04501/2013), and FCT project PTDC/DTP_PIC/5587/2014 at the University of Aveiro, Portugal. Lausanne study: This work was supported by grants from the Swiss National Research Foundation (SNF 320030_141179). PAGES study: Harald Hampel is an employee of Eisai Inc. During part of this work he was supported by the AXA Research Fund, the “Fondation partenariale Sorbonne Université” and the “Fondation pour la Recherche sur Alzheimer”, Paris, France. Mannheim, Germany Biobank: Department of geriatric Psychiatry, Central Institute for Mental Health, Mannheim, University of Heidelberg, Germany. Genotyping for the Swedish Twin Studies of Aging was supported by NIH/NIA grant R01 AG037985. Genotyping in TwinGene was supported by NIH/NIDDK U01 DK066134. WvdF is recipient of Joint Programming for Neurodegenerative Diseases (JPND) grants PERADES (ANR-13-JPRF-0001) and EADB (733051061). Gothenburg Birth Cohort (GBC) Studies: We would like to thank UCL Genomics for performing the genotyping analyses. The studies were supported by The Stena Foundation, The Swedish Research Council (2015-02830, 2013-8717), The Swedish Research Council for Health, Working Life and Wellfare (2013-1202, 2005-0762, 2008-1210, 2013-2300, 2013- 2496, 2013-0475), The Brain Foundation, Sahlgrenska University Hospital (ALF), The Alzheimer’s Association (IIRG-03-6168), The Alzheimer’s Association Zenith Award (ZEN-01-3151), Eivind och Elsa K:son Sylvans Stiftelse, The Swedish Alzheimer Foundation. Clinical AD, Sweden: We would like to thank UCL Genomics for performing the genotyping analyses. Barcelona Brain Biobank: Brain Donors of the Neurological Tissue Bank of the Biobanc-Hospital Clinic-IDIBAPS and their families for their generosity. Hospital Clínic de Barcelona Spanish Ministry of Economy and Competitiveness-Instituto de Salud Carlos III and Fondo Europeo de Desarrollo Regional (FEDER), Unión Europea, “Una manera de hacer Europa” grants (PI16/0235 to Dr. R. Sánchez-Valle and PI17/00670 to Dr. A.Antonelli). AA is funded by Departament de Salut de la Generalitat de Catalunya, PERIS 2016-2020 (SLT002/16/00329). Work at JP-T laboratory was possible thanks to funding from Ciberned and generous gifts from Consuelo Cervera Yuste and Juan Manuel Moreno Cervera. Sydney Memory and Ageing Study (Sydney MAS): We gratefully acknowledge and thank the following for their contributions to Sydney MAS: participants, their supporters and the Sydney MAS Research Team (current and former staff and students). Funding was awarded from the Australian National Health and Medical Research Council (NHMRC) Program Grants (350833, 568969, 109308). AddNeuroMed consortium was led by Simon Lovestone, Bruno Vellas, Patrizia Mecocci, Magda Tsolaki, Iwona Kłoszewska, Hilkka Soininen. This work was supported by InnoMed (Innovative Medicines in Europe), an integrated project funded by the European Union of the Sixth Framework program priority (FP6-2004- LIFESCIHEALTH-5). Oviedo: This work was partly supported by Grant from Fondo de Investigaciones Sanitarias-Fondos FEDER EuropeanUnion to Victoria Alvarez PI15/00878. Project MinE: The ProjectMinE study was supported by the ALS Foundation Netherlands and the MND association (UK) (Project MinE, www.projectmine.com). The SPIN cohort: We are indebted to patients and their families for their participation in the “Sant Pau Initiative on Neurodegeneration cohort”, at the Sant Pau Hospital (Barcelona). This is a multimodal research cohort for biomarker discovery and validation that is partially funded by Generalitat de Catalunya (2017 SGR 547 to JC), as well as from the Institute of Health Carlos III-Subdirección General de Evaluación and the Fondo Europeo de Desarrollo Regional (FEDER- “Una manera de Hacer Europa”) (grants PI11/02526, PI14/01126, and PI17/01019 to JF; PI17/01895 to AL), and the Centro de Investigación Biomédica en Red Enfermedades Neurodegenerativas programme (Program 1, Alzheimer Disease to AL). We would also like to thank the Fundació Bancària Obra Social La Caixa (DABNI project) to JF and AL; and Fundación BBVA (to AL), for their support in funding this follow-up study. Adolfo López de Munain is supported by Fundación Salud 2000 (PI2013156), CIBERNED and Diputación Foral de Gipuzkoa (Exp.114/17). Pascual Sánchez-Juan is supported by CIBERNED and Carlos III Institute of Health, Spain (PI08/0139, PI12/02288, and PI16/01652, PI20/01011), jointly funded by Fondo Europeo de Desarrollo Regional (FEDER), Unión Europea, “Una manera de hacer Europa”. We thank Biobanco Valdecilla for their support. Amsterdam dementia Cohort (ADC): Research of the Alzheimer center Amsterdam is part of the neurodegeneration research program of Amsterdam Neuroscience. The AlzheimerCenter Amsterdam is supported by Stichting Alzheimer Nederland and Stichting VUmc fonds. The clinical database structure was developed with funding from Stichting Dioraphte. Genotyping of the Dutch case-control samples was performed in the context of EADB (European Alzheimer&Dementia biobank) funded by the JPco-fuND FP-829-029 (ZonMW project number #733051061). This research is performed by using data from the Parelsnoer Institute an initiative of the Dutch Federation of University Medical Centres (www.parelsnoer.org). 100- Plus study: We are grateful for the collaborative efforts of all participating centenarians and their family members and/or relations. We thank the Netherlands Brain Bank for supplying DNA for genotyping. This work was supported by Stichting AlzheimerNederland (WE09.2014-03), Stichting Diorapthe, Horstingstuit foundation, Memorabel (ZonMW project number #733050814, #733050512) and Stichting VUmcFonds. Additional support for EADB cohorts: WF, SL, HH are recipients of ABOARD, a public-private partnership receiving funding from ZonMW (#73305095007) and Health∼Holland, Topsector Life Sciences & Health (PPP-allowance; #LSHM20106). The DELCODE study was funded by the German Center for Neurodegenerative Diseases (Deutsches Zentrum für Neurodegenerative Erkrankungen (DZNE)), reference number BN012.

Gra@ce. The Genome Research @ Fundació ACE project (GR@ACE) is supported by Grifols SA, Fundación bancaria ‘La Caixa’, Fundació ACE, and CIBERNED (Centro de Investigación Biomédica en Red Enfermedades Neurodegenerativas (Program 1, Alzheimer Disease to MB and AR)). A.R. and M.B. receive support from the European Union/EFPIA Innovative Medicines Initiative Joint undertaking ADAPTED and MOPEAD projects (grant numbers 115975 and 115985, respectively). M.B. and A.R. are also supported by national grants PI13/02434, PI16/01861, PI17/01474 and PI19/01240. Acción Estratégica en Salud is integrated into the Spanish National R + D + I Plan and funded by ISCIII (Instituto de Salud Carlos III)–Subdirección General de Evaluación and the Fondo Europeo de Desarrollo Regional (FEDER–’Una manera de hacer Europa’). Some control samples and data from patients included in this study were provided in part by the National DNA Bank Carlos III (www.bancoadn.org, University of Salamanca, Spain) and Hospital Universitario Virgen de Valme (Sevilla, Spain); they were processed following standard operating procedures with the appropriate approval of the Ethical and Scientific Committee. The present work has been performed as part of the doctoral program of I. de Rojas at the Universitat de Barcelona (Barcelona, Spain).

EADI. This work has been developed and supported by the LABEX (laboratory of excellence program investment for the future) DISTALZ grant (Development of Innovative Strategies for a Transdisciplinary approach to ALZheimer’s disease) including funding from MEL (Metropole européenne de Lille), ERDF (European Regional Development Fund) and Conseil Régional Nord Pas de Calais. This work was supported by INSERM, the National Foundation for Alzheimer’s disease and related disorders, the Institut Pasteur de Lille and the Centre National de Recherche en Génomique Humaine, CEA, the JPND PERADES, the Laboratory of Excellence GENMED (Medical Genomics) grant no. ANR-10-LABX-0013 managed by the National Research Agency (ANR) part of the Investment for the Future program, and the FP7 AgedBrainSysBio. The Three-City Study was performed as part of collaboration between the Institut National de la Santé et de la Recherche Médicale (Inserm), the Victor Segalen Bordeaux II University and Sanofi-Synthélabo. The Fondation pour la Recherche Médicale funded the preparation and initiation of the study. The 3C Study was also funded by the Caisse Nationale Maladie des Travailleurs Salariés, Direction Générale de la Santé, MGEN, Institut de la Longévité, Agence Française de Sécurité Sanitaire des Produits de Santé, the Aquitaine and Bourgogne Regional Councils, Agence Nationale de la Recherche, ANR supported the COGINUT and COVADIS projects. Fondation de France and the joint French Ministry of Research/INSERM “Cohortes et collections de données biologiques” programme. Lille Génopôle received an unconditional grant from Eisai. The Three-city biological bank was developed and maintained by the laboratory for genomic analysis LAG-BRC - Institut Pasteur de Lille.

Rotterdam Study. The Rotterdam Study is funded by Erasmus Medical Center and Erasmus University, Rotterdam, Netherlands Organization for the Health Research and Development (ZonMw), the Research Institute for Diseases in the Elderly (RIDE), the Ministry of Education, Culture and Science, the Ministry for Health, Welfare and Sports, the European Commission (DG XII), and the Municipality of Rotterdam. The authors are grateful to the study participants, the staff from the Rotterdam Study and the participating general practitioners and pharmacists. The generation and management of GWAS genotype data for the Rotterdam Study (RS-I, RS-II, RSIII) was executed by the Human Genotyping Facility of the Genetic Laboratory of the Department of Internal Medicine, Erasmus MC, Rotterdam, The Netherlands. The GWAS datasets are supported by the Netherlands Organization of Scientific Research NOW Investments (Project number 175.010.2005.011, 911-03-012), the Genetic Laboratory of the Department of Internal Medicine, Erasmus MC, the Research Institute for Diseases in the Elderly (014- 93-015; RIDE2), the Netherlands Genomics Initiative (NGI)/Netherlands Organization for Scientific Research (NWO) Netherlands Consortium for Healthy Aging (NCHA), project number 050-060-810.

ADSP. The Alzheimer’s Disease Sequencing Project (ADSP) is comprised of two Alzheimer’s Disease (AD) genetics consortia and three National Human Genome Research Institute (NHGRI) funded Large Scale Sequencing and Analysis Centers (LSAC). The two AD genetics consortia are the Alzheimer’s Disease Genetics Consortium (ADGC) funded by NIA (U01 AG032984), and the Cohorts for Heart and Aging Research in Genomic Epidemiology (CHARGE) funded by NIA (R01 AG033193), the National Heart, Lung, and Blood Institute (NHLBI), other National Institute of Health (NIH) institutes and other foreign governmental and non-governmental organizations. The Discovery Phase analysis of sequence data is supported through UF1AG047133 (to Drs. Schellenberg, Farrer, Pericak-Vance, Mayeux, and Haines); U01AG049505 to Dr. Seshadri; U01AG049506 to Dr. Boerwinkle; U01AG049507 to Dr. Wijsman; and U01AG049508 to Dr. Goate and the Discovery Extension Phase analysis is supported through U01AG052411 to Dr. Goate, U01AG052410 to Dr. Pericak-Vance and U01 AG052409 to Drs. Seshadri and Fornage.

The ADGC cohorts included in ADSP include: Adult Changes in Thought (ACT) (UO1 AG006781, UO1 HG004610, UO1 HG006375, U01 HG008657), the Alzheimer’s Disease Centers (ADC) (P30 AG019610, P30 AG013846, P50 AG008702, P50 AG025688, P50 AG047266, P30 AG010133, P50 AG005146, P50 AG005134, P50 AG016574, P50 AG005138, P30 AG008051, P30 AG013854, P30 AG008017, P30 AG010161, P50 AG047366, P30 AG010129, P50 AG016573, P50 AG016570, P50 AG005131, P50 AG023501, P30 AG035982, P30 AG028383, P30 AG010124, P50 AG005133, P50 AG005142, P30 AG012300, P50 AG005136, P50 AG033514, P50 AG005681, and P50 AG047270), the Chicago Health and Aging Project (CHAP) (R01 AG11101, RC4 AG039085, K23 AG030944), Indianapolis Ibadan (R01 AG009956, P30 AG010133), the Memory and Aging Project (MAP) (R01 AG17917), Mayo Clinic (MAYO) (R01 AG032990, U01 AG046139, R01 NS080820, RF1 AG051504, P50 AG016574), Mayo Parkinson’s Disease controls (NS039764, NS071674, 5RC2HG005605), University of Miami (R01 AG027944, R01 AG028786, R01 AG019085, IIRG09133827, A2011048), the Multi-Institutional Research in Alzheimer’s Genetic Epidemiology Study (MIRAGE) (R01 AG09029, R01 AG025259), the National Cell Repository for Alzheimer’s Disease (NCRAD) (U24 AG21886), the National Institute on Aging Late Onset Alzheimer’s Disease Family Study (NIA- LOAD) (R01 AG041797), the Religious Orders Study (ROS) (P30 AG10161, R01 AG15819), the Texas Alzheimer’s Research and Care Consortium (TARCC) (funded by the Darrell K Royal Texas Alzheimer’s Initiative), Vanderbilt University/Case Western Reserve University (VAN/CWRU) (R01 AG019757, R01 AG021547, R01 AG027944, R01 AG028786, P01 NS026630, and Alzheimer’s Association), the Washington Heights-Inwood Columbia Aging Project (WHICAP) (RF1 AG054023), the University of Washington Families (VA Research Merit Grant, NIA: P50AG005136, R01AG041797, NINDS: R01NS069719), the Columbia University Hispanic Estudio Familiar de Influencia Genetica de Alzheimer (EFIGA) (RF1 AG015473), the University of Toronto (UT) (funded by Wellcome Trust, Medical Research Council, Canadian Institutes of Health Research), and Genetic Differences (GD) (R01 AG007584). The CHARGE cohorts are supported in part by National Heart, Lung, and Blood Institute (NHLBI) infrastructure grant HL105756 (Psaty), RC2HL102419 (Boerwinkle) and the neurology working group is supported by the National Institute on Aging (NIA) R01 grant AG033193.

The CHARGE cohorts participating in the ADSP include the following: Austrian Stroke Prevention Study (ASPS), ASPS-Family study, and the Prospective Dementia Registry-Austria (ASPS/PRODEM-Aus), the Atherosclerosis Risk in Communities (ARIC) Study, the Cardiovascular Health Study (CHS), the Erasmus Rucphen Family Study (ERF), the Framingham Heart Study (FHS), and the Rotterdam Study (RS). ASPS is funded by the Austrian Science Fond (FWF) grant number P20545-P05 and P13180 and the Medical University of Graz. The ASPS-Fam is funded by the Austrian Science Fund (FWF) project I904), the EU Joint Programme - Neurodegenerative Disease Research (JPND) in frame of the BRIDGET project (Austria, Ministry of Science) and the Medical University of Graz and the Steiermärkische Krankenanstalten Gesellschaft. PRODEM-Austria is supported by the Austrian Research Promotion agency (FFG) (Project No. 827462) and by the Austrian National Bank (Anniversary Fund, project 15435. ARIC research is carried out as a collaborative study supported by NHLBI contracts (HHSN268201100005C, HHSN268201100006C, HHSN268201100007C, HHSN268201100008C, HHSN268201100009C, HHSN268201100010C, HHSN268201100011C, and HHSN268201100012C). Neurocognitive data in ARIC is collected by U01 2U01HL096812, 2U01HL096814, 2U01HL096899, 2U01HL096902, 2U01HL096917 from the NIH (NHLBI, NINDS, NIA and NIDCD), and with previous brain MRI examinations funded by R01-HL70825 from the NHLBI. CHS research was supported by contracts HHSN268201200036C, HHSN268200800007C, N01HC55222, N01HC85079, N01HC85080, N01HC85081, N01HC85082, N01HC85083, N01HC85086, and grants U01HL080295 and U01HL130114 from the NHLBI with additional contribution from the National Institute of Neurological Disorders and Stroke (NINDS). Additional support was provided by R01AG023629, R01AG15928, and R01AG20098 from the NIA. FHS research is supported by NHLBI contracts N01-HC-25195 and HHSN268201500001I. This study was also supported by additional grants from the NIA (R01s AG054076, AG049607 and AG033040 and NINDS (R01 NS017950). The ERF study as a part of EUROSPAN (European Special Populations Research Network) was supported by European Commission FP6 STRP grant number 018947 (LSHG-CT-2006- 01947) and also received funding from the European Community’s Seventh Framework Programme (FP7/2007-2013)/grant agreement HEALTH-F4- 2007-201413 by the European Commission under the programme “Quality of Life and Management of the Living Resources” of 5th Framework Programme (no. QLG2-CT-2002- 01254). High-throughput analysis of the ERF data was supported by a joint grant from the Netherlands Organization for Scientific Research and the Russian Foundation for Basic Research (NWO-RFBR 047.017.043). The Rotterdam Study is funded by Erasmus Medical Center and Erasmus University, Rotterdam, the Netherlands Organization for Health Research and Development (ZonMw), the Research Institute for Diseases in the Elderly (RIDE), the Ministry of Education, Culture and Science, the Ministry for Health, Welfare and Sports, the European Commission (DG XII), and the municipality of Rotterdam. Genetic data sets are also supported by the Netherlands Organization of Scientific Research NWO Investments (175.010.2005.011, 911-03-012), the Genetic Laboratory of the Department of Internal Medicine, Erasmus MC, the Research Institute for Diseases in the Elderly (014- 93-015; RIDE2), and the Netherlands Genomics Initiative (NGI)/Netherlands Organization for Scientific Research (NWO) Netherlands Consortium for Healthy Aging (NCHA), project 050-060-810. All studies are grateful to their participants, faculty and staff. The content of these manuscripts is solely the responsibility of the authors and does not necessarily represent the official views of the National Institutes of Health or the U.S. Department of Health and Human Services.

The FUS cohorts include: the Alzheimer’s Disease Centers (ADC) (P30 AG019610, P30 AG013846, P50 AG008702, P50 AG025688, P50 AG047266, P30 AG010133, P50 AG005146, P50 AG005134, P50 AG016574, P50 AG005138, P30 AG008051, P30 AG013854, P30 AG008017, P30 AG010161, P50 AG047366, P30 AG010129, P50 AG016573, P50 AG016570, P50 AG005131, P50 AG023501, P30 AG035982, P30 AG028383, P30 AG010124, P50 AG005133, P50 AG005142, P30 AG012300, P50 AG005136, P50 AG033514, P50 AG005681, and P50 AG047270), Alzheimer’s Disease Neuroimaging Initiative (ADNI) (U19AG024904), Amish Protective Variant Study (RF1AG058066), Cache County Study (R01AG11380, R01AG031272, R01AG21136, RF1AG054052), Case Western Reserve University Brain Bank (CWRUBB) (P50AG008012), Case Western Reserve University Rapid Decline (CWRURD) (RF1AG058267, NU38CK000480), CubanAmerican Alzheimer’s Disease Initiative (CuAADI) (3U01AG052410), Estudio Familiar de Influencia Genetica en Alzheimer (EFIGA) (5R37AG015473, RF1AG015473, R56AG051876), Genetic and Environmental Risk Factors for Alzheimer Disease Among African Americans Study (GenerAAtions) (2R01AG09029, R01AG025259, 2R01AG048927), Gwangju Alzheimer and Related Dementias Study (GARD) (U01AG062602), Hussman Institute for Human Genomics Brain Bank (HIHGBB) (R01AG027944, Alzheimer’s Association “Identification of Rare Variants in Alzheimer Disease”), Ibadan Study of Aging (IBADAN) (5R01AG009956), Mexican Health and Aging Study (MHAS) (R01AG018016), Multi-Institutional Research in Alzheimer’s Genetic Epidemiology (MIRAGE) (2R01AG09029, R01AG025259, 2R01AG048927), Northern Manhattan Study (NOMAS) (R01NS29993), Peru Alzheimer’s Disease Initiative (PeADI) (RF1AG054074), Puerto Rican 1066 (PR1066) (Wellcome Trust (GR066133/GR080002), European Research Council (340755)), Puerto Rican Alzheimer Disease Initiative (PRADI) (RF1AG054074), Reasons for Geographic and Racial Differences in Stroke (REGARDS) (U01NS041588), Research in African American Alzheimer Disease Initiative (REAAADI) (U01AG052410), Rush Alzheimer’s Disease Center (ROSMAP) (P30AG10161, R01AG15819, R01AG17919), University of Miami Brain Endowment Bank (MBB), and University of Miami/Case Western/North Carolina A&T African American (UM/CASE/NCAT) (U01AG052410, R01AG028786). The four LSACs are: the Human Genome Sequencing Center at the Baylor College of Medicine (U54 HG003273), the Broad Institute Genome Center (U54HG003067), The American Genome Center at the Uniformed Services University of the Health Sciences (U01AG057659), and the Washington University Genome Institute (U54HG003079).

