## Supplementary material for "Challenges at the *APOE* locus: A robust quality control approach for accurate *APOE* genotyping"

### Supplemental Methods

#### Phenotype Ascertainment

*Cohorts and Phenotype Ascertainment*

In the current study, we used data from twenty-nine cohorts and three sequencing projects related to AD[1-23]. Details on phenotype ascertainment are described elsewhere[1–6]. Briefly, all individuals with a diagnosis of AD met National Institute of Neurological and Communicative Disorders and Stroke/Alzheimer's Disease and Related Disorders Association (NINCDS-ADRDA) criteria for probable or possible late onset AD[7], or met Diagnosis and Statistical Manual of Mental Disorders IV-V (DSMIV-V) criteria[8–10], or had a clinical dementia rating (CDR® Dementia Staging Instrument[11]) > 0.5. Some cohorts verified AD diagnoses by means of neuropathology, using Braak staging[12], CERAD scoring[13], or National Institute on Aging Reagan (NIA-Reagan) 1997 criteria[14]. Cognitively normal subjects (controls) did not have AD according to the above clinical criteria for AD, did not have a diagnosis of MCI, and had a CDR of 0 and/or Mini-Mental State Examination (MMSE[15]) > 25. In the MIRAGE cohort, control status was evaluated through a Modified Telephone Interview of Cognitive Status score ≥ 86 (a telephone version of the MMSE)[16].

Further, the National Alzheimer's Coordinating Center (NACC), Rush University Religious Orders Study/Memory and Aging Project (ROSMAP), and Alzheimer's Disease Neuroimaging Initiative (ADNI), are longitudinal cohorts that provide detailed information regarding clinical status (control, MCI, demented) and presumed disease etiology at repeated examinations. Additionally, deceased subjects are assessed for neuropathology. Where possible, in NACC, a final diagnoses of MCI or possible/probable/definite AD was obtained using NIA Alzheimer's Association (NIA-AA) 2011 criteria[17,18]. In all three cohorts, AD diagnoses were verified by neuropathology as middle or high AD likelihood following NIA-Reagan 1997 criteria (moderate to frequent neuritic plaques and Braak stage III-VI)[14]. In concordance with the category "possible AD dementia with evidence of the AD pathophysiological process" from the NIA-AA 2011 criteria[17], we attributed possible AD diagnoses to subjects who met clinical criteria for non-AD dementia but also met AD neuropathological criteria. In concordance with the NIA-AA 2011/2012 framework[18,19], we also evaluated neuropathology in MCI subjects to verify presumed AD etiology (cf. page 5). Controls were not re-evaluated based on neuropathology data. Subjects that reverted from dementia to control status during longitudinal follow-up were excluded. Additional cohort-specific details are listed below.

*NACC*

Genotyping waves 1 through 7 from the Alzheimer's Disease Centers (ADC1-7) and a subset of the ADSP projects include subjects ascertained and evaluated by the clinical and neuropathological cores of 32 NIA-funded ADCs. NACC coordinates the collection of these phenotypes, implements diagnoses (cognitively normal, cognitively impaired but not MCI, MCI, demented; and presumed disease etiology) and then provides all data to researchers under the form of the Minimum Data Set (MDS), Uniform Data Set (UDS)[20–22], and Neuropathology data set (NP)[23]. The MDS represents an older subset of the NACC data and only contains cross-sectional data, while the more recent UDS provides longitudinal phenotypes and covariates. Since 2015, the UDS was updated to incorporate the NIA-AA 2011 criteria for MCI and AD[18,24]. In the current study, we used the UDS and NP for which data was collected between September 2005 and March 2020. The ADC1-7 data sets covered 7,627 subjects in the UDS and 2,629 subjects in the MDS.

Subjects that had a diagnosis of Down syndrome, central nervous system neoplasm, bipolar disorder, schizophrenia, alcohol-induced dementia, or substance-abuse-induced dementia, were excluded. Subjects carrying mutations of dominantly inherited AD or frontotemporal lobar degeneration (FTLD) were also excluded. Subjects with a final diagnosis of MCI or dementia, for which the etiology was unknown, not due to AD, or only secondary due to AD (and without AD neuropathological information), were excluded. Subjects with a final diagnosis of "cognitively impaired but not MCI", but having no other neurological disorder, were kept as controls, considering that this more consistently matched control criteria in many of the other cohorts considered in this study.

*ROSMAP*

In ROSMAP, subjects were diagnosed at each visit: as possible/probable AD according to NINCDS-ADRDA criteria[7]; as MCI when judged to have cognitive impairment but not meeting dementia criteria according to the clinician; or as control when there was no cognitive impairment or the subject did not meet dementia criteria[25,26]. At time of death, a final clinical diagnosis was made by an expert neurologist, followed by case conference consensus review (blinded to postmortem data)[27].

*ADNI*

In ADNI, subjects were diagnosed at regular visits: as possible/probable AD according to NINCDS-ADRDA criteria[7]; as MCI according to Petersen/Winblad criteria; or as control when not demented, not MCI, CDR = 0, and MMSE > 28. Neuropathology assessments followed the NACC NP framework.

*Discovery Samples Phenotype Harmonization*

The discovery sample contained many subjects that were genotyped multiple times across different studies. This largely reflected efforts from the ADGC, ADSP, and AMP-AD, to perform next generation sequencing (NGS) on existing cohort samples for the purpose of rare variant discovery and AD gene prioritization. In other instances, participants were recruited in different studies at different times. Therefore, to handle potential duplicate discordance and phenotype heterogeneity, we implemented a cross-sample phenotype harmonization procedure aiming to standardize pathology-verified diagnoses where possible, share unique missing information across all duplicate entries of a given subject, resolve longitudinal changes in diagnosis, and flag subjects with unresolvable duplicate discordance for exclusion.

Duplicate samples were identified by determining genetic cryptic relatedness (cf. page 6 below), but for the purpose of sample cross-referencing did not include known identical twins in LOAD and ROSMAP samples. First, duplicate samples were flagged as discordant if their age-at-death information differed by more than 2 years or if pathology measures (Braak or neuritic plaque density) differed. Across all cohorts, where possible, AD diagnoses were verified by neuropathology as middle or high AD likelihood following NIA-Reagan 1997 criteria (moderate to frequent neuritic plaques and Braak stage III-VI)[14]. Additionally, when only either neuritic plaque or Braak information was available and in line with NIA-Reagan 1997 middle or high AD likelihood criteria, and/or the cohort/project demographics provided a diagnosis of definite AD, the subject was considered to have pathology-verified AD status. Cognitively normal (CN) subjects with evidence of AD pathology were kept as CN. Further, if at least one entry across duplicate samples indicated a diagnosis of Down syndrome, central nervous system neoplasm, bipolar disorder, schizophrenia, alcohol-induced dementia, substance-abuse-induced dementia, neurological (not including Parkinson’s disease) or systemic disease despite being cognitively normal, or carrying mutations of dominantly inherited AD or frontotemporal lobar degeneration (FTLD), then all duplicate samples were marked as such and flagged for exclusion. Extending on the above, all genetic samples were checked for the presence of known pathogenic mutations on *APP, PSEN1, PSEN2* and *MAPT*, whereby carriers and their duplicate samples were flagged for exclusion.

Then, duplicate samples with differing age entries (i.e. longitudinal changes) were evaluated. Reversions from AD or dementia to MCI status, or from MCI to cognitively normal (CN) status, were permitted, but reversions from AD or non-AD dementia to CN status were flagged for exclusion. “Reversions” from AD to non-AD dementia status were permitted, unless pathology (cf. above) indicated the presence of AD pathology, thereby marking the subject as AD. Vice versa, “conversions” from non-AD dementia to AD status were permitted, unless pathology (cf. above) indicated no presence of AD pathology, thereby marking the subject as non-AD dementia. All other types of conversions were directly permitted. Then, duplicate samples for which the diagnoses at the oldest shared age entries differed, or for which diagnoses differed but age was consistent (i.e. apparent cross-sectional discordances), were evaluated. Discordances between AD and non-AD dementia status were resolved on the basis of pathology (cf. above) or flagged as discordant if no pathology data was available. Discordances between CN and AD status, or CN and non-AD dementia status, were resolved as respectively AD or non-AD dementia when those dementia diagnoses corresponded to a unique age-at-onset (of symptoms) without other available age information (i.e. indicating that a conversion likely occurred after the subject was lost to follow-up in the cohort that last observed a CN status), or, were flagged as discordant if duplicate entries shared the same age-at-examination and age-at-last-exam. Discordances between CN and MCI status, or MCI and AD status, or MCI and non-AD dementia status, were resolved as respectively MCI, AD, or non-AD dementia (i.e. keeping the most severe diagnosis).

Finally, once all clinical diagnostic and pathological data were unified across duplicate entries, pathological criteria were applied once more to obtain the final diagnoses. Where possible, AD diagnoses were verified by neuropathology as middle or high AD likelihood following NIA-Reagan 1997 criteria (moderate to frequent neuritic plaques and Braak stage III-VI)[14]. In concordance with the category "possible AD dementia with evidence of the AD pathophysiological process" from the NIA-AA 2011 criteria[17], we attributed possible AD diagnoses to subjects who met clinical criteria for non-AD dementia but also met AD neuropathological criteria. In concordance with the NIA-AA 2011/2012 framework[18,19], we also evaluated neuropathology in MCI subjects to verify presumed AD etiology and considered subjects as cases if AD pathology, following NIA-Reagan 1997 criteria (cf. above), was present (i.e. marking high likelihood of AD etiology). Controls were not re-evaluated based on neuropathology data.

Beyond cross-referencing clinical diagnostic and pathological data across subjects, other covariates were considered for cross-referencing or sharing in case of missingness across duplicate entries. These included age-at-onset of cognitive symptoms, age-at-examination providing clinical diagnosis, at-at-last exam, age-at-death, sex, race, ethnicity, *APOE* genotype provided from demographics, *APOE* genotype provided from whole-genome sequencing, and *APOE* genotype provided from whole-exome sequencing. Duplicate entries with discordant sex or race information were flagged for exclusion.

### Genetic Data Quality Control and Processing

*Discovery Samples Genetic Data Harmonization and Standard Quality Control*

Genotypes were available from commercial high-density single-nucleotide polymorphism (SNP) genotyping microarrays (Illumina or Affymetrix), Whole-exome sequencing (WES), or Whole-genome sequencing (WGS) (**Table S1**). Genotype samples had their genetic variants lifted to hg19 using liftOver if not released in hg19[28]. Autosomal variants were extracted from the SNP array data and further processed in several stages. First, SNP array data were processed by the Genotype Harmonizer with CEU and TSI HapMap populations as the reference panel, to perform automatic strand alignment[29]. Then, multi-allelic SNPs, SNPs located on common copy number or segmental duplication regions, and duplicated or monomorphic SNPs, were removed. The list of multi-allelic SNPs or SNPs located on common copy number and segmental duplication regions was created using Tri-Typer[30]. The list of CNV and segmental duplication regions was curated from the Eichler lab (eichlerlab.gs.washington.edu/database.html)[31] and the gnomAD website (gnomad.broadinstitute.org/downloads)[32]. All respective genotype data sets were then iteratively merged with each other, applying strand flipping and variant ID updating as applicable, to ultimately obtain parsimonious data sets that could be merged for cross-sample relationship determination and principal component analyses (cf. below).

Genetic data were then further processed using Plink v1.9. The numbers of remaining samples after each quality control (QC) or processing step are listed in **Table S2**. For each sample platform, subjects with autosome missingness (≥ 5%) and sex problems (discordance between genetic sex and demographic sex, or deviation of expected X-chromosome homozygosity/heterozygosity) were flagged for exclusion.

*Discovery Samples Ancestry Determination*

Individual ancestries were determined using SNPweights v.2.1 with populations from the 1000 Genomes Consortium as a reference[33,34]. By applying an ancestry percentage cut-off ≥ 75%, the samples were stratified into the five super populations, South-Asians (SAS), East-Asians (EAS), Americans (AMR), Africans (AFR) and Europeans (EUR) (**Figure S1**). Subjects with a genetic ancestry that differed from their race, as provided in cohort demographics, were flagged for exclusion.

*Discovery Samples Relationship Determination using Plink*

Across all cohorts (and ethnicities) the relatedness of subjects (after QC indicated above) was evaluated through identity-by-descent (IBD) analysis (using directly genotyped non-palindromic SNPs that were shared across all genetic datasets with a call rate > 99%, minor allele frequency (MAF) > 1%). This IBD outcome (**Figure S3-A**) was used for duplicate (IBD > 0.95) tracking across samples.

*Discovery Samples Relationship Determination and Principal Component Analysis using GENESIS*

Across SNP array and WGS data (or SNP array, WES, and WGS data for duplicate identification), the relatedness of subjects and principal components capturing population substructure were determined using IBD and principal component analyses (PCA) as implemented through the R package GENESIS (R v3.6.0)[35]. Specifically, this approach first uses an R-implementation of KING-robust to determine kinship coefficients that take into account ancestry divergence. The derived pairwise kinship coefficients are then used to perform a PCA in related samples (PC-AiR) providing accurate ancestry inference not confounded by family structure. The latter output is then used to estimate kinship coefficients using PC-Relate, which accounts for population structure (ancestry) among sample individuals through the use of ancestry representative principal components (PCs) to provide accurate relatedness estimates due only to recent family (pedigree) structure. For each respective data merge, these analyses were performed on directly genotyped and pruned SNPs ($R^2 < 0.5$, call rate > 99.9%, MAF > 1%, and excluding palindromic SNPs) in non-Hispanic white European ancestry individuals (**Figure S2-3**).

*Discovery Samples APOE genotype assessment in old and new ADSP WES/WGS*

The “old” ASDP WES (N=10,919) and ADSP WGS (N=4,750), which were available to us prior to 2020, refer to the ADSP discovery and extension phases described in detail in the main text. These data were used for variant association analyses. The new ADSP WES (N=20,503) and ADSP WGS (N=16,906), which were available to us as of 2021, refer to updated samples (including the prior ADSP data and more) and updated joint calling performed by the ADSP (NG00067.v5)[36]. These data were used to verify *APOE* genotypes (and filter discordant subjects) in variant association analyses for approach 2.

In both the old and new ADSP WGS, rs429358 and rs7412 showed low genotype missingness across subjects, reflecting good variant quality metrics in the joint calling performed by ADSP. In the old and new ADSP WES, rs7412 respectively did not pass ADSP quality control and showed a high genotype missingness at (32.5%). This resulted from a low read depth and genotype quality in some of the different WES capture kits that were used in the ADSP WES[2]. We therefore sought to re-call both variants in order to fill out missing *APOE* information where possible. We first inferred the variants' genotype using data called by the ADSP, which required a read depth read depth (DP) >=10 and genotype quality (GQ) >= 20. We then further inferred the variants' genotype if DP and GQ were respectively greater than or equal to 6 and 20, observing at least 20% alternate allele reads to call a heterozygote (e.g. *APOE**3/4).

After this first round of *APOE* genotype ascertainment, some individuals still had either the rs7412 or rs429358 genotype missing (i.e., only one of the two variants could be called using the above criteria), making it impossible to infer their *APOE* genotype from the ADSP NGS data alone. Many of these remaining individuals however had a reported *APOE* genotype in their demographics that could be used to complete the missing information in a second additional round of *APOE* genotype ascertainment. This approach was preferred over relying solely on the *APOE* genotype in the demographics, since the genotype calls on the ADSP NGS data are expected to provide higher accuracy compared to other commonly used *APOE* direct genotyping methods[37]. To illustrate, consider the example where one of these remaining individuals in the sequencing data was homozygous for the reference allele at rs429358, which would suggest the subject is *APOE**3/3, but had a missing genotype at rs7412. In this case, from the ADSP NGS data, we know that this individual is not carrying an *APOE**4 allele, but we cannot determine the presence or absence of an *APOE**2 allele. We then turned to the information from the *APOE* genotype provided in the demographics to infer the most likely *APOE* genotype. For the current example, if the individual has a provided *APOE* genotype that was 2/2, 2/3, or 3/3, then the information in the ADSP NGS data is deemed concordant with the provided *APOE* genotype (that is, rs429358 is always the reference allele for those provided *APOE* genotypes) and we used the provided *APOE* genotype. However, if the provided *APOE* genotype was 4/4 or 3/4, then we would correct it to *APOE**3/3, because the ADSP NGS information clearly indicated there was no *APOE**4 genotype call (similarly a provided *APOE**2/4 genotype would be corrected to *APOE**2/3). This can be generalized as: for remaining individuals with DP>=6 and GQ>=20 at rs429358, the ADSP NGS data at rs429358 was used to change, when discordant, the provided *APOE**3 genotype to *APOE**4, or vice-versa. One additional extension to this step was implemented for the few scenarios where the ADSP NGS data called two rs429358 alleles (i.e. *APOE**4/4) but the allelic distribution indicated that the reference allele was still observed (e.g. 1 REF allele and 7 ALT alleles). In these situations, if the provided *APOE* genotype indicated the presence of *APOE**3, then the genotype was corrected to *APOE**3/4 (reasoning there is sufficient evidence to support the presence of an *APOE**3 genotype). The extra checks described in this paragraph were also applied to subjects in the first QC round (prior paragraph), who had 6<=DP<10 and GQ>=20 for both rs429358 and rs7412.

As a quality check, using these thresholds, we did not observe any discordance in the inferred *APOE* genotype across 3,499 duplicates between the ADSP WGS and ADSP WES.

**Statistical Analyses – Additional Model Criteria**

*Case-Control Analyses*

For age adjustment, when multiple age data were available, we prioritized in cases age-at-onset of symptoms (AAO) > age-at-examination providing clinical diagnosis of AD (AAE) > age-at-death (AAD), and in controls AAD > age-at-last-examination (AAL) (**Table S3**). This priority ranking is consistent with prior AD studies[1,38]. For cases that only had AAD available, the final ages used for regression analysis were subtracted by 10 years in order to approximate AAO. This reflects expected mean delays between AAO and AAD for AD patients[39], and is consistent with the derived age covariate for AD case-control analyses provided by the Alzheimer's Disease Genetics Consortium (ADGC) on NIAGADS[40]. In the ADNI and ROSMAP cohorts, which provide longitudinal diagnoses with matching age information, but do not directly provide AAO, the AAE variable used either the age-at-MCI-diagnosis or the first age-at-dementia-diagnosis.

*Models including unrelated individuals*

In sensitivity analyses that did not include related individuals, only a single subject was retained per relatedness cluster, prioritizing first younger cases followed by older controls.

**Figure S1. Admixture plot for the five major super populations across all discovery samples.** Black vertical line marks the cut-off for EUR ancestry [>=75%].

*Abbreviations: EUR, European; AFR, African; AMR; American; SAS, Southern Asian; EAS; Eastern Asian.*

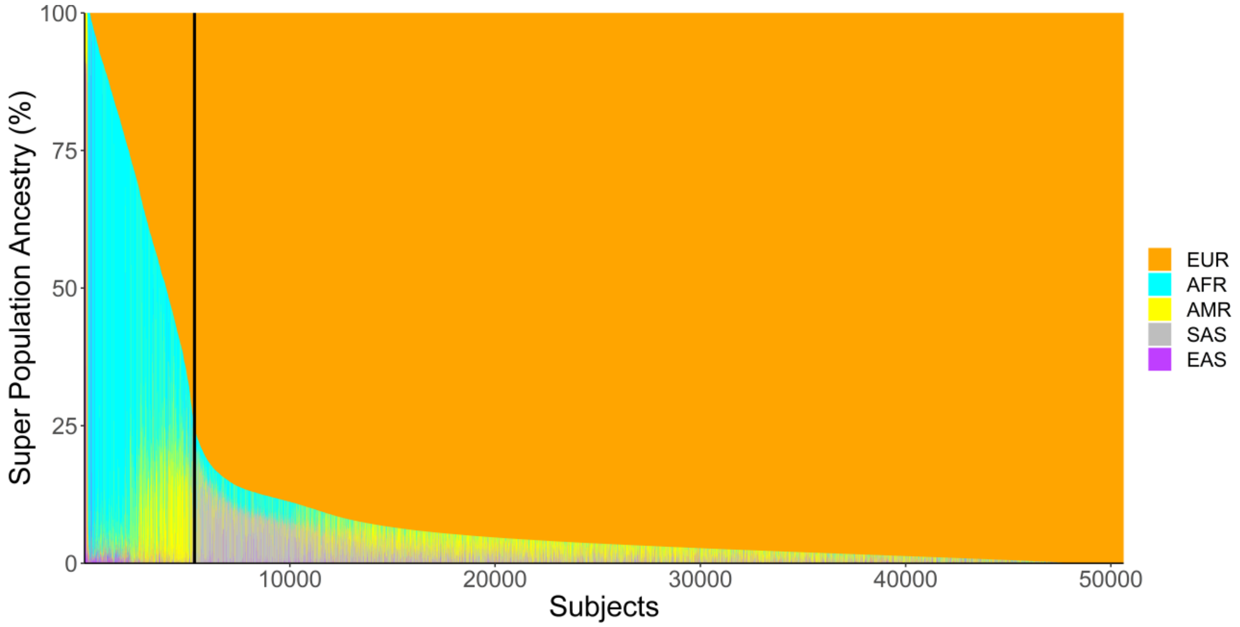

**Figure S2. First five principal components of the genetic population structure in European subjects from the discovery samples. (A)** PCs are labelled by sub-European ancestries for merged SNP array and WGS data.

*Abbreviations: PC, principal component; EU, European; NWE, Northwestern European; SEE, Southeastern European; AJE, Ashkenazi Jewish.*

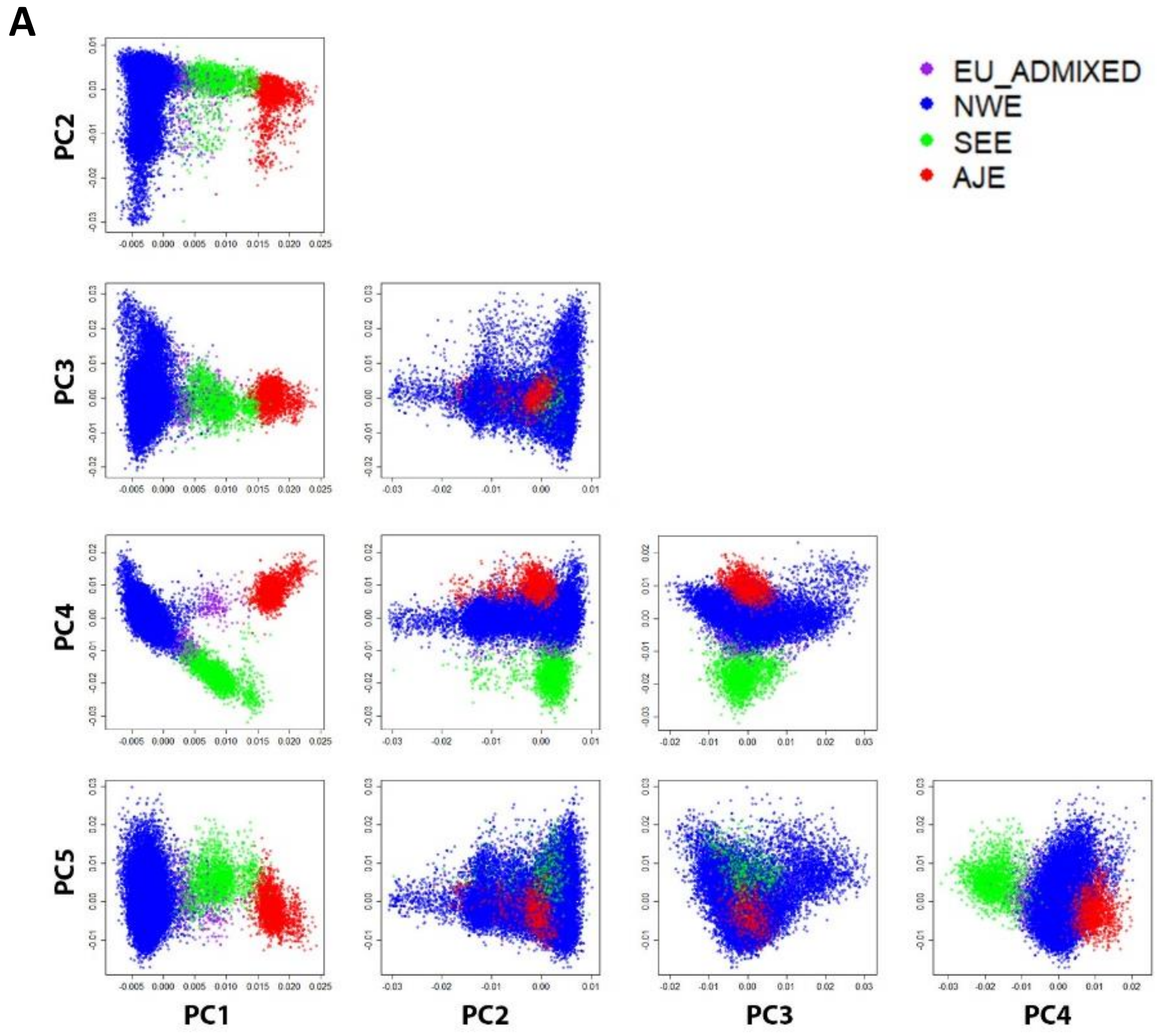

**Figure S2. (B)** PCs are labelled by genetic source for merged SNP array and WGS data.

*Abbreviations: PC, principal component; SNP, single nucleotide polymorphism; WGS, whole genome sequencing; ADSP, Alzheimer's Disease Sequencing Project.*

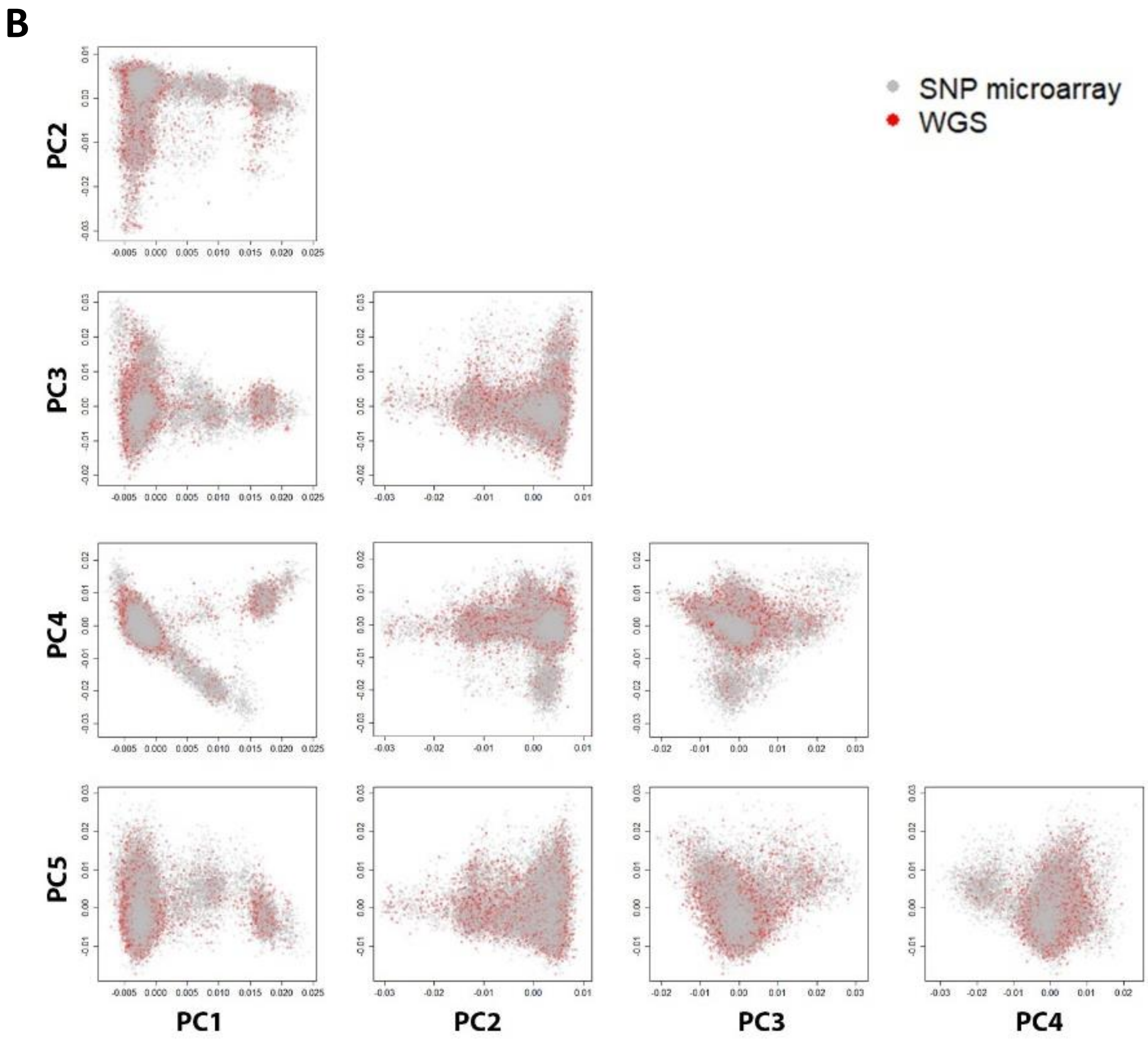

**Figure S2. (C)** PCs are labelled by Diagnosis. The Diagnosis of "Other" refers to subjects with diagnoses such as "demented not due to AD" or "mild cognitive impairment".

*Abbreviations: PC, principal component; CN, Cognitively normal; AD, Alzheimer's disease.*

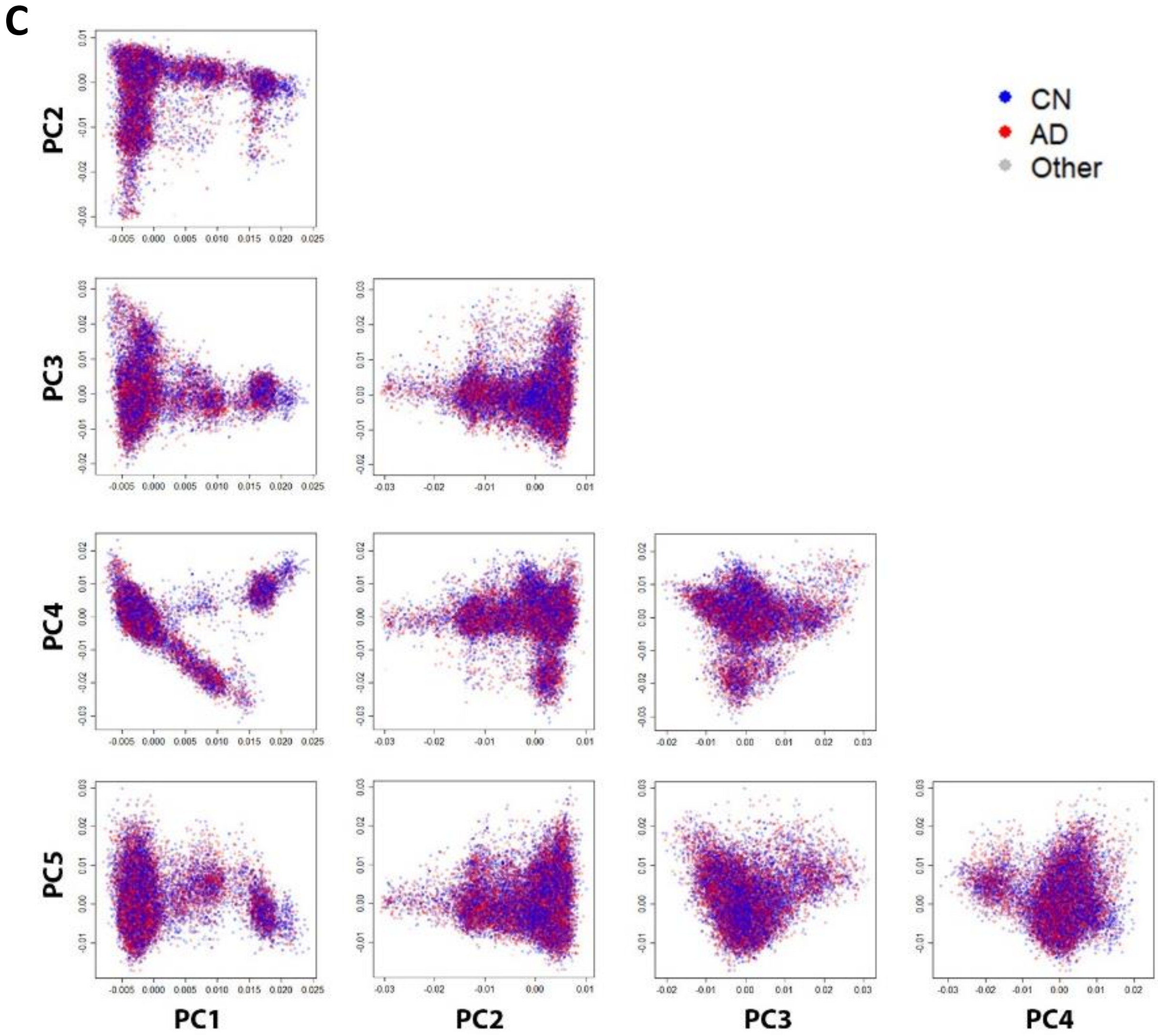

**Figure S3. Identity-by-descent (IBD) analyses.** Panel shows scatter a density plot, with values ranging from 0 (blue) to 200 or more (red) IBD pairs. Dashed lines indicate boundaries between levels of relatedness. All IBD analyses were thresholded at the lower boundary of 4th degree relatedness (using KING-robust and PC-Air/PC-Relate, the expected reliable lower boundary for dense SNP data is 3rd or 4th degree relatedness[35]). The panel shows an IBD analyses performed on a merge of SNP array and WGS data using KING-robust and PC-Air/PC-Relate. X-axis shows k0, indicating the probability of having 0 alleles identical by descent (ranging 0 to 1). Y-axis shows kin, the pairwise kinship coefficient estimate (ranging 0 to 0.5 conventionally, but possibly extending beyond these boundaries given the use of ancestry divergence measures by KING and PC-Relate). Note the well-conditioned relationship of k0 and kin down to the lower boundary of 3rd degree relatedness. High intensity inflation was visible at the lower boundary of 4th degree relatedness, suggesting inflated number of IBD pairs. Relatedness levels in this approach were thus judged to be reasonably reliable down to 3rd degree relatedness.

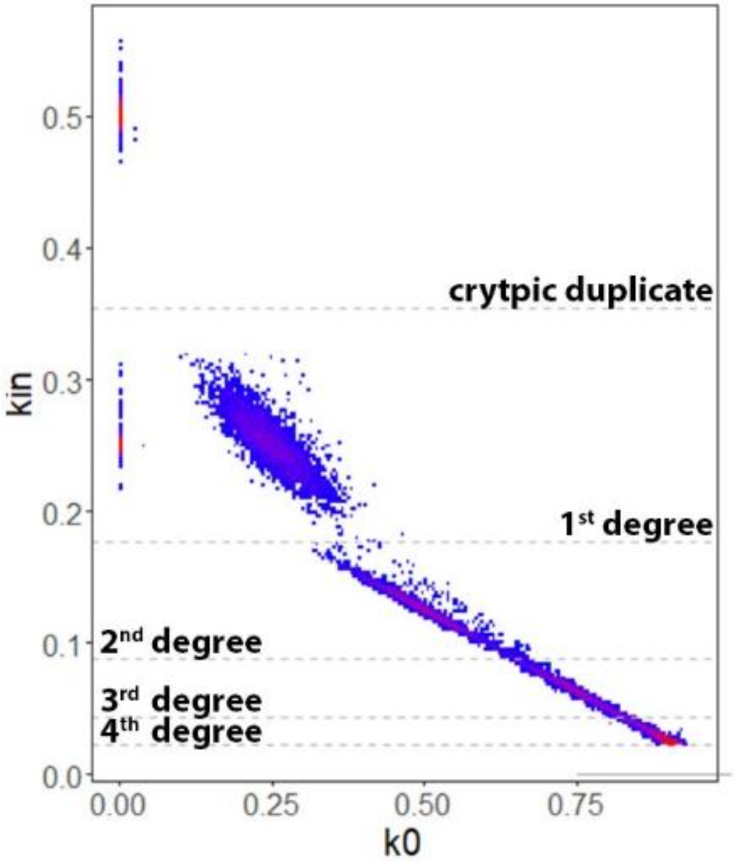

**Figure S4. Genotype intensity data in EADB for rs439401.** Top panel shows full sample, bottom panel shows *APOE**4/4 subjects.

*Abbreviations: GT, genotype.*

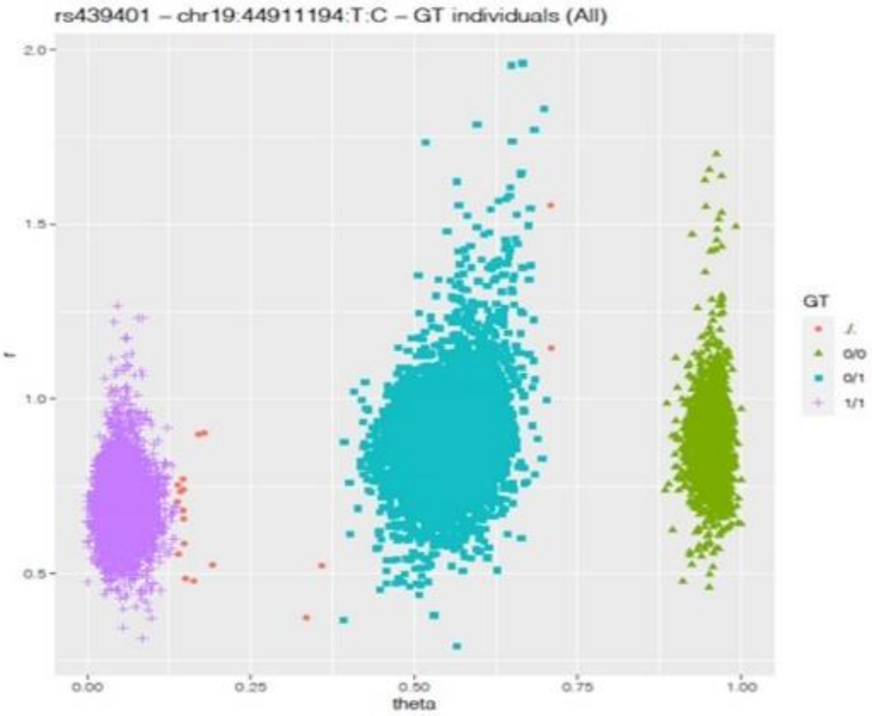

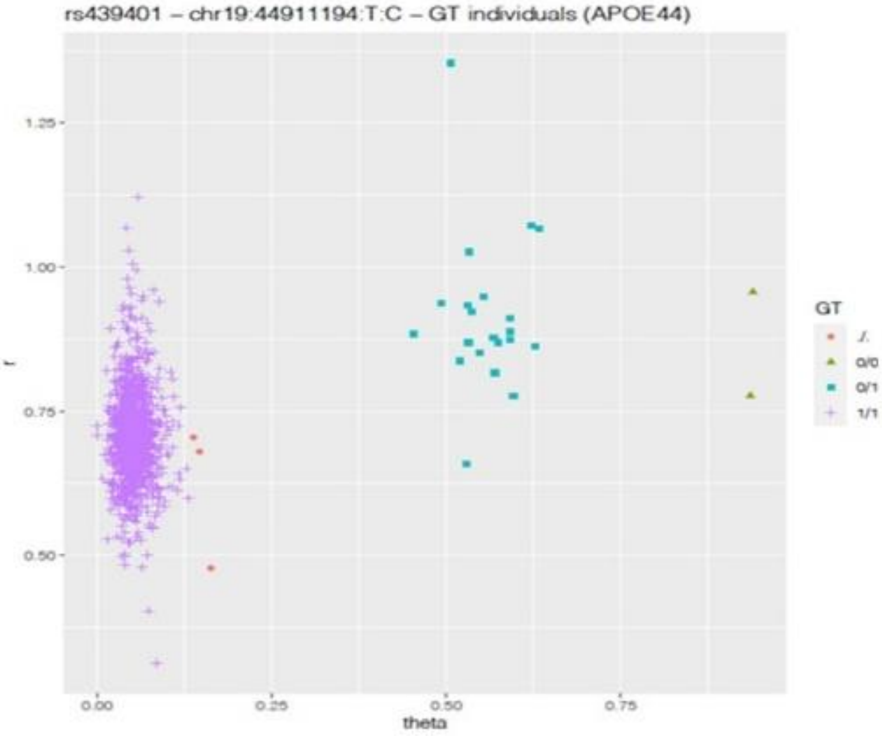

**Figure S5. Discovery case-control association results for rs439401 when in-phase with *APOE**4, comparing the basic model to cohort and cohort/array/center adjustment, using *APOE* filtering approach 1.**

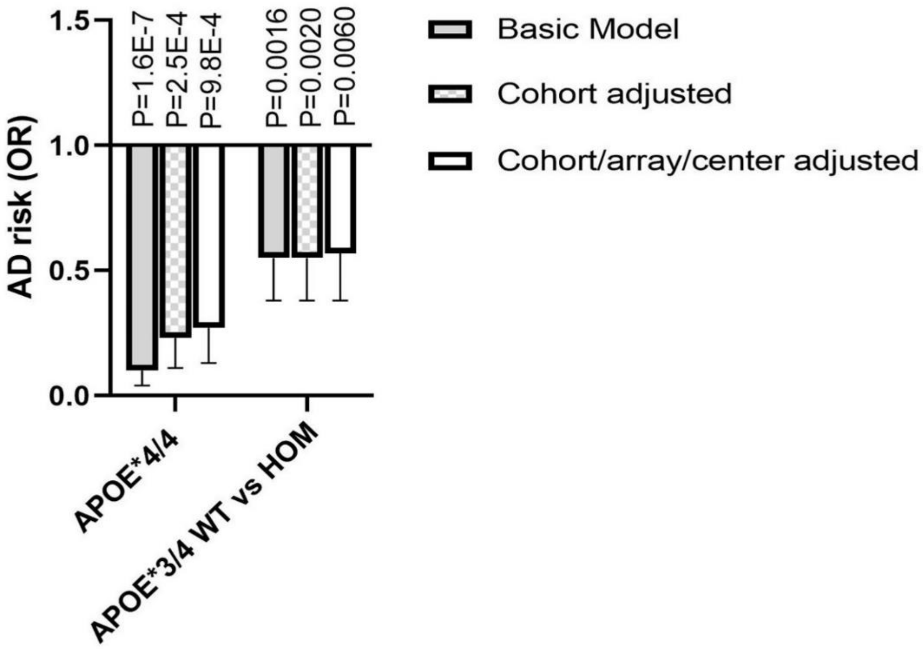

**Figure S6. Simulation of the concordance between observed *APOE**4/4 and true *APOE**4/4 genotypes considering different type I and II error rates. A)** Controls. **B)** Cases. Four different true frequencies were considered for controls and cases respectively. The frequencies were centered around the frequency observed in the full discovery cohort, which is somewhere around the expected true frequency. Type I error rate is defined as the probability to mis-classify non-*APOE**4/4 as *APOE**4/4; Type II error rate is defined as the probability to mis-classify *APOE**4/4 as non-*APOE**4/4.

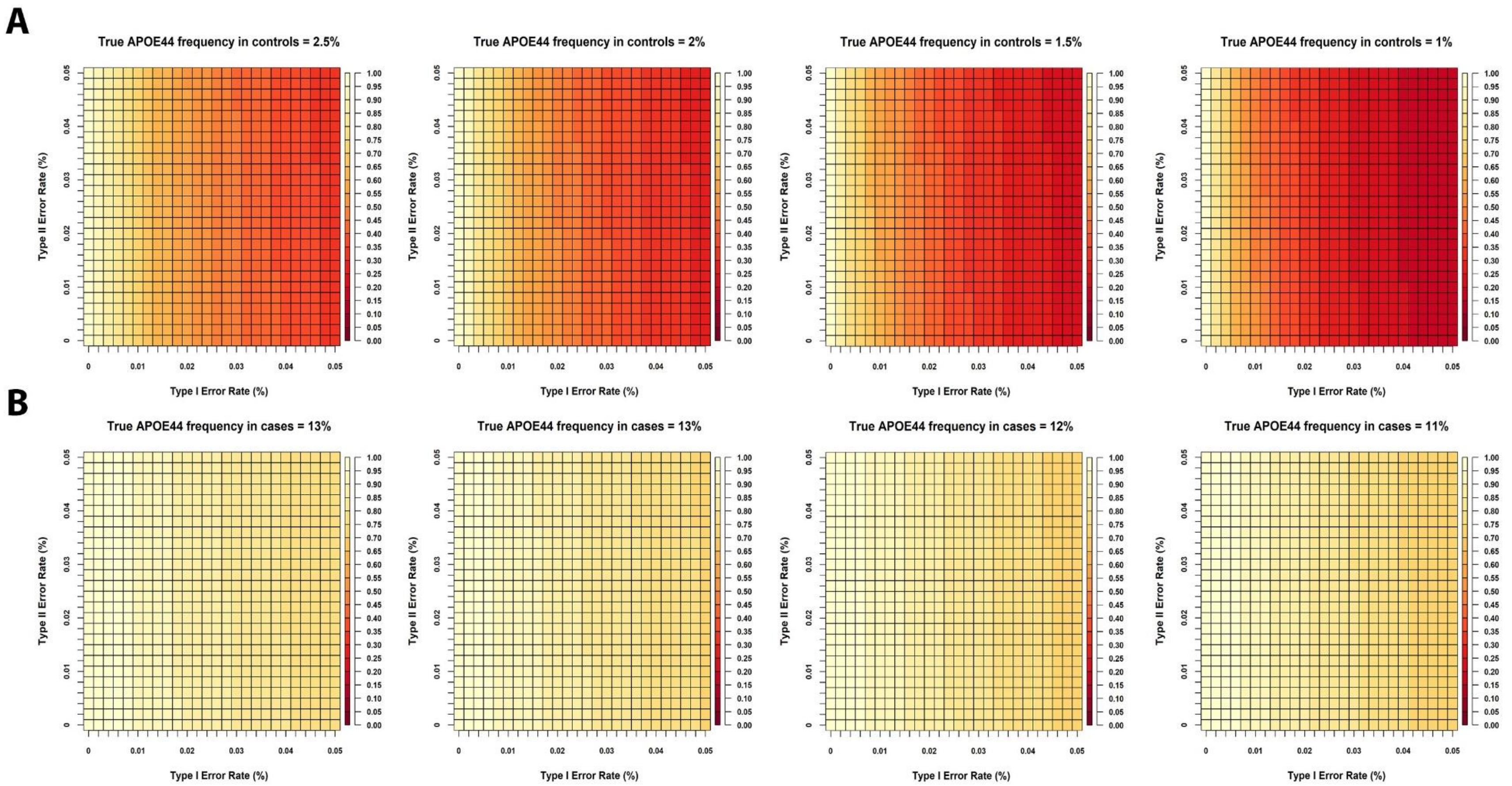

**Figure S7. Simulation of the concordance between observed *APOE**4/4 and true *APOE**4/4 genotypes considering different type I and II error rates and considering only *APOE**3/4 and *APOE**4/4 carriers. A)** Controls. **B)** Cases. One limitation when considering concordance rates as depicted in Figure S9 is that this is assumes each *APOE* genotype (2/2, 2/3, 3/3, 2/4, 3/4, 4/4) could equally produce any other *APOE* genotype when error occurs. Based on observations in Table S18, it may be more likely that *APOE**4/4 subjects are enriched for miscalled *APOE**3/4 subjects. Observed versus true *APOE**4/4 concordance rates may thus be better assessed by considering the true frequency of APOE*4/4 subjects with regard to the group of *APOE**3/4 and *APOE**4/4 subjects.

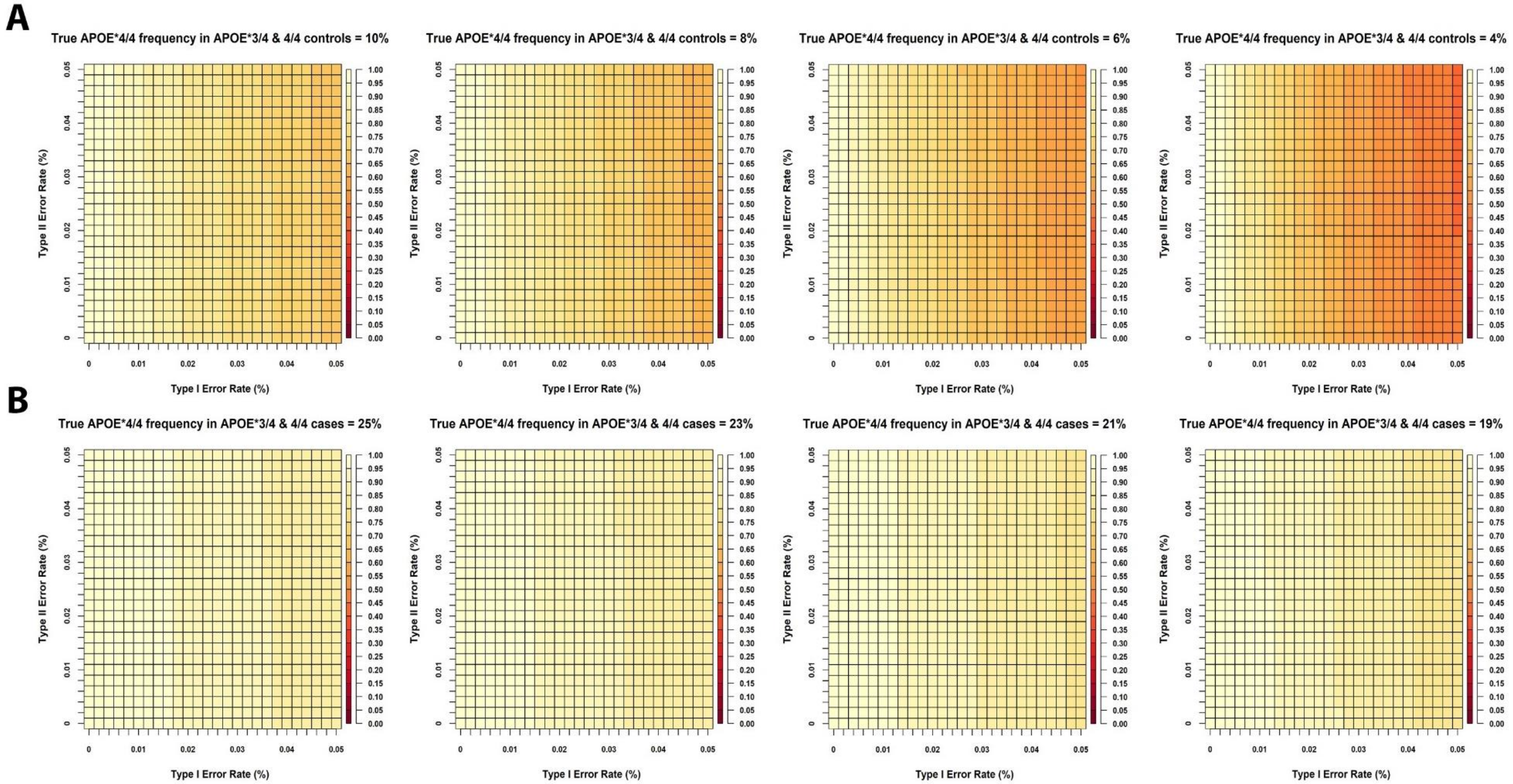

**Figure S8. Concordance rates between imputed and provided *APOE* across different *APOE* strata. A)** Combining cases and controls per stratum. **B)** Splitting cases and controls per stratum. Considered samples were those used in the discovery analyses, passing sample QC, and here further excluding subjects that did not have imputed genotypes available. Further, the analyses considered the provided *APOE* genotype to stratify samples as depicted (not using verification from the WGS/WES *APOE* genotypes). Number of subjects across groups are shown at the top of bar graphs. *Abbrevations: CN, Cognitively normal; AD, Alzheimer's Disease.*

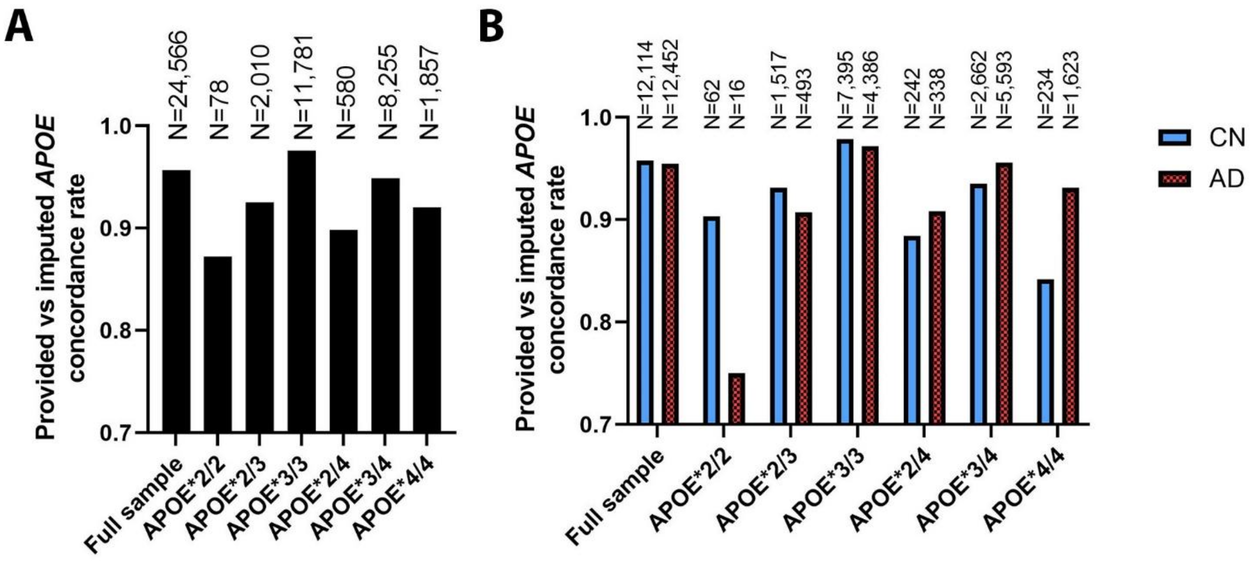

**Table S1. Overview genotyping platforms across cohorts/projects considered in analysis approach 1.**

| Cohort/Project | Genotyping Platform | In-house data ID | Sample count | Data Repository & ID |
|---|---|---|---|---|
| ACT | Illumina Human 660W-Quad | ACT | 2790 | NIAGADS (NG00034) / dbGaP (phs000234) |
| ADC1 | Illumina Human 660W-Quad | ADC1 | 2731 | NIAGADS (NG00022) / NACC |
| ADC2 | Illumina Human 660W-Quad | ADC2 | 928 | NIAGADS (NG00023) / NACC |
| ADC3 | Illumina Human OmniExpress | ADC3 | 1526 | NIAGADS (NG00024) / NACC |
| ADC4 | Illumina Human OmniExpress | ADC4 | 1054 | NIAGADS (NG00068) / NACC |
| ADC5 | Illumina Human OmniExpress | ADC5 | 1224 | NIAGADS (NG00069) / NACC |
| ADC6 | Illumina Human OmniExpress | ADC6 | 1333 | NIAGADS (NG00070) / NACC |
| ADC7 | Illumina Infinium Human OmniExpressExome | ADC7 | 1462 | NIAGADS (NG00071) / NACC |
| ADDNEUROMED | Illumina Human 610-Quad | ADM_Q | 315 | Synapse AddNeuroMed (syn4907804) |
| | Illumina Human OmniExpress | ADM_O | 329 | Synapse AddNeuroMed (syn4907804) |
| ADNI | Illumina Human 610-Quad | ADNI_1 | 757 | LONI ADNI |
| | Illumina Human OmniExpress | ADNI_2 | 361 | LONI ADNI |
| | Illumina Omni 2.5 | ADNI_O25 | 812 | LONI ADNI |
| | Whole Genome Sequencing - Illumina | ADNI_WGS | 812 | LONI ADNI |
| ADNI-DOD | Illumina Human OmniExpress | ADNI_DOD | 204 | LONI ADNIDOD |
| ADSP discovery | Whole Exome Sequencing (discovery joint called) - Baylor College | ADSP_DISC_Baylor | 2609 | dbGaP (phs000572) / NACC |
| | Whole Exome Sequencing (discovery joint called) - Broad Institute | ADSP_DISC_Broad | 4574 | dbGaP (phs000572) / NACC |
| | Whole Exome Sequencing (discovery joint called) - Washington University | ADSP_DISC_Washu | 3726 | dbGaP (phs000572) / NACC |
| | Whole Exome Sequencing (discovery joint called) - unspecified | ADSP_DISC | 10 | dbGaP (phs000572) / NACC |
| ADSP extension | Whole Genome Sequencing (extension joint called) - Baylor College | ADSP_EXT_Baylor | 1260 | NIAGADS (NG00067) / NACC |
| | Whole Genome Sequencing (extension joint called) - Broad Institute | ADSP_EXT_Broad | 1365 | NIAGADS (NG00067) / NACC |
| | Whole Genome Sequencing (extension joint called) - Illumina | ADSP_EXT_Illumina | 809 | NIAGADS (NG00067) / NACC |
| | Whole Genome Sequencing (extension joint called) - Washington University | ADSP_EXT_Washu | 1316 | NIAGADS (NG00067) / NACC |
| EADI | Illumina Human 610-Quad | EADI | 9863 | contact: |
| EADB | Illumina Global Screening Array | EADB | 64704 | contact: |

| | | | | |
|---|---|---|---|---|
| GenADA | Affymetrix 500K | GSK | 1571 | dbGaP (phs000219) |
| NIA-LOAD | Illumina Human 610-Quad | LOAD | 5220 | NIAGADS (NG00020) |
| MAYO | Illumina Human Hap300 | MAYO_1 | 2099 | Synapse AMP-AD (syn5591675) / NIAGADS (NG00029) |
| | Whole Genome Sequencing (AMP-AD joint called ROSMAP/MAYO/MSBB) | AMP_AD_MAYO_WGS | 349 | Synapse AMP-AD (syn22264775) |
| MAYO2 | Illumina Omni 2.5 | MAYO_2 | 314 | Synapse AMP-AD (syn5550404) |
| | Whole Genome Sequencing (AMP-AD joint called ROSMAP/MAYO/MSBB) | AMP_AD_MAYO_WGS | 349 | Synapse AMP-AD (syn22264775) |
| MIRAGE | Illumina Human CNV370-Duo | MIRAGE_370 | 397 | NIAGADS (NG00031) |
| | Illumina Human 610-Quad | MIRAGE_610 | 1105 | NIAGADS (NG00031) |
| MSBB | Whole Genome Sequencing (AMP-AD joint called ROSMAP/MAYO/MSBB) | AMP_AD_MSBB_WGS | 349 | Synapse AMP-AD (syn3159438, syn22264775) |
| MTC | Illumina Human OmniExpress | MTC | 542 | NIAGADS (NG00096) |
| OHSU | Illumina Human CNV370-Duo | OHSU | 647 | NIAGADS (NG00017) |
| ROTTERDAM | Illumina Infinium II HumanHap550chip v3.0 | ROTTERDAM | 10150 | contact: |
| ROSMAP | Affymetrix GeneChip 6.0 - Broad Institute | ROSMAP_1B | 1126 | RADC Rush (contact:) / Synapse AMP-AD (syn3219045) |
| | Affymetrix GeneChip 6.0 - TGen | ROSMAP_1T | 582 | RADC Rush (contact:) / Synapse AMP-AD (syn3219045) |
| | Illumina Human OmniExpress 12 - Chop | ROSMAP_2C | 382 | RADC Rush (contact:) / Synapse AMP-AD (syn7824841) |
| | Illumina Multi-Ethnic - BU | ROSMAP_3BU | 494 | RADC Rush (contact:) |
| | Whole Genome Sequencing (AMP-AD joint called ROSMAP/MAYO/MSBB) | AMP_AD_ROSMAP_WGS | 1196 | RADC Rush (contact:) / Synapse AMP-AD (syn22264775) |
| TARCC | Affymetrix 6.0 | TARCC | 625 | NIAGADS (NG00097) |
| TGEN2 | Affymetrix 6.0 | TGEN | 1599 | NIAGADS (NG00028) |
| UPITT | Illumina Human Omni1-Quad | UPITT | 2440 | NIAGADS (NG00026) |
| UM/VU/MSSM | Illumina Human 1M-Duo, Illumina 1M | UVM_A | 1153 | NIAGADS (NG00042) |
| | Affymetrix 6.0 | UVM_B | 864 | NIAGADS (NG00042) |
| | Illumina Human 550K. Illumina Human 610-Quad | UVM_C | 445 | NIAGADS (NG00042) |
| WASHU | Illumina Human 610-Quad | WASHU_1 | 670 | NIAGADS (NG00030) |
| WASHU2 | Illumina Human OmniExpress | WASHU_2 | 235 | NIAGADS (NG00087) |
| WHICAP | Illumina Human OmniExpress | WHICAP | 647 | NIAGADS (NG00093) |

33

**Table S2. Discovery sample sizes per cohort/project after sequential quality control and filtering steps (detailed in column titles).** Cohorts/Arrays that had no coverage of rs439401 (genotyping rate 0%) are not listed.

| Cohorts | 1. All genotyped subjects | 2. Genotype missingness <95% | 3. No sex problems/missingness | 4. No ancestry/race discordance | 5. No duplicate discordance | 6. Filter to CN/AD | 7. *APOE* genotype available | 8. AGE available | 9. AGE 60y and up | 10. European (EU) | 11. Retain unique non-duplicate |
|---|---|---|---|---|---|---|---|---|---|---|---|
| ACT | 2790 | 2790 | 2790 | 2786 | 2784 | 2558 | 2364 | 2364 | 2363 | 2270 | 2269 |
| ADC1 | 2731 | 2731 | 2730 | 2697 | 2682 | 2522 | 2508 | 2508 | 2221 | 2058 | 2053 |
| ADC2 | 928 | 928 | 927 | 927 | 923 | 829 | 827 | 827 | 820 | 817 | 816 |
| ADC3 | 1526 | 1526 | 1525 | 1524 | 1519 | 1356 | 1354 | 1354 | 1253 | 1232 | 1232 |
| ADC4 | 1054 | 1054 | 1054 | 1054 | 1054 | 875 | 871 | 871 | 839 | 828 | 827 |
| ADC5 | 1224 | 1224 | 1223 | 1222 | 1221 | 1005 | 1005 | 1005 | 996 | 993 | 989 |
| ADC6 | 1333 | 1333 | 1332 | 1332 | 1330 | 933 | 927 | 927 | 719 | 719 | 716 |
| ADC7 | 1462 | 1462 | 1462 | 1460 | 1460 | 1314 | 1314 | 1314 | 1313 | 1308 | 1308 |
| ADM_O | 315 | 315 | 313 | 313 | 313 | 240 | 235 | 235 | 232 | 231 | 231 |
| ADM_Q | 329 | 329 | 281 | 281 | 281 | 202 | 201 | 201 | 196 | 196 | 196 |
| ADNI_1 | 757 | 756 | 755 | 751 | 750 | 551 | 549 | 549 | 538 | 495 | 293 |
| ADNI_2 | 361 | 361 | 359 | 353 | 353 | 248 | 248 | 248 | 241 | 206 | 185 |
| ADNI_O25 | 812 | 812 | 811 | 804 | 804 | 470 | 470 | 470 | 462 | 433 | 362 |
| ADNI_DOD | 204 | 204 | 203 | 200 | 200 | 94 | 94 | 94 | 94 | 79 | 79 |
| ADSP_EXT_Baylor | 1260 | 1259 | 1259 | 1253 | 1251 | 1064 | 1064 | 1064 | 1033 | 119 | 119 |
| ADSP_EXT_Broad | 1365 | 1362 | 1362 | 1356 | 1352 | 1274 | 1274 | 1274 | 1245 | 972 | 291 |
| ADSP_EXT_Illumina | 809 | 809 | 808 | 801 | 801 | 468 | 468 | 468 | 460 | 432 | 1 |
| ADSP_EXT_WashU | 1316 | 1316 | 1315 | 1301 | 1298 | 1248 | 1248 | 1247 | 1204 | 144 | 32 |
| LOAD | 5220 | 5220 | 5201 | 5164 | 5155 | 4482 | 4470 | 4470 | 3940 | 3344 | 3050 |
| MAYO_1 | 2099 | 2058 | 2058 | 2058 | 2056 | 1953 | 1918 | 1918 | 1917 | 1887 | 1761 |
| MAYO_2 | 314 | 314 | 312 | 312 | 311 | 223 | 223 | 223 | 210 | 208 | 185 |
| AMP_AD_MAYO_WGS | 349 | 349 | 349 | 349 | 348 | 263 | 263 | 263 | 250 | 250 | 45 |
| MIRAGE_370 | 397 | 397 | 358 | 358 | 358 | 356 | 331 | 329 | 321 | 295 | 276 |
| MIRAGE_610 | 1105 | 1105 | 992 | 991 | 990 | 984 | 926 | 915 | 901 | 857 | 822 |
| AMP_AD_MSBB_WGS | 349 | 349 | 340 | 337 | 334 | 224 | 224 | 224 | 216 | 186 | 180 |
| MTC | 542 | 542 | 540 | 539 | 539 | 460 | 371 | 367 | 365 | 352 | 332 |
| OHSU | 647 | 641 | 634 | 634 | 618 | 576 | 561 | 561 | 559 | 554 | 511 |
| ROSMAP_2C | 382 | 382 | 380 | 380 | 380 | 298 | 298 | 298 | 298 | 293 | 261 |
| ROSMAP_3BU | 494 | 488 | 480 | 452 | 449 | 353 | 303 | 303 | 303 | 231 | 180 |
| AMP_AD_ROSMAP_WGS | 1196 | 1196 | 1185 | 1184 | 1167 | 937 | 937 | 937 | 937 | 937 | 647 |
| UPITT | 2440 | 2371 | 2364 | 2364 | 2364 | 2235 | 2227 | 2218 | 2218 | 2204 | 2163 |
| UVM_A | 1153 | 1153 | 1153 | 1153 | 1151 | 1147 | 1140 | 1140 | 1140 | 1137 | 1099 |
| UVM_B | 864 | 864 | 863 | 863 | 861 | 816 | 804 | 800 | 795 | 791 | 553 |
| WASHU | 670 | 670 | 668 | 668 | 667 | 640 | 629 | 599 | 582 | 582 | 434 |
| WHICAP | 647 | 647 | 647 | 647 | 647 | 639 | 638 | 638 | 638 | 631 | 622 |
| Total | 39444 | 39317 | 39033 | 38868 | 38771 | 33837 | 33284 | 33223 | 31819 | 28271 | 25120 |

**Table S3. Expanded sample demographics for Alzheimer's disease case-control association analyses.**

| Cohort | | Diagnosis | | Pathology | | Sex | Age | Age type | | | |
|---|---|---|---|---|---|---|---|---|---|---|---|
| Name | Participants after QC (N) | Type | (N) | Available (N (%)) | AD Path. (N (%)) | Female (N (%)) | Age (Mean (SD)) | AAD (Mean (SD)) [%] | AAL (Mean (SD)) [%] | AAE (Mean (SD)) [%] | AAO (Mean (SD)) [%] |
| **DISCOVERY SAMPLE** | | | | | | | | | | | |
| **ACT** | 2269 | CN | 1645 | 196 (11.9 %) | 0 (0.0 %) | 891 (54.2 %) | 83.5 (5.9) | 83.7 (5.7) [59.4 %] | 83.2 (6.3) [40.6 %] | - | - |
| | | AD | 624 | 119 (19.1 %) | 119 (100 %) | 405 (64.9 %) | 81.8 (6.3) | - | - | 82.6 (5.8) [7.9 %] | 81.8 (6.3) [92.1 %] |
| **NACC - ADC1** | 2053 | CN | 490 | 152 (31.0 %) | 8 (5.3 %) | 279 (56.9 %) | 81.4 (9.1) | 85.8 (8.5) [39.2 %] | 78.6 (8.3) [60.8 %] | - | - |
| | | AD | 1563 | 1535 (98.2 %) | 1535 (100 %) | 857 (54.8 %) | 73.0 (7.5) | 83.5 (6.0) [1.6 %] | - | 79.6 (8.5) [6.3 %] | 72.6 (7.2) [92.1 %] |
| **NACC - ADC2** | 816 | CN | 125 | 18 (14.4 %) | 4 (22.2 %) | 88 (70.4 %) | 79.8 (9.1) | 86.3 (8.3) [21.6 %] | 78.0 (8.6) [78.4 %] | - | - |
| | | AD | 691 | 329 (47.6 %) | 329 (100 %) | 362 (52.4 %) | 73.1 (7.0) | - | - | 76.9 (6.9) [1.3 %] | 73.1 (7.0) [98.7 %] |
| **NACC - ADC3** | 1232 | CN | 490 | 42 (8.6 %) | 5 (11.9 %) | 305 (62.2 %) | 79.3 (9.5) | 88.3 (7.5) [20.4 %] | 77.0 (8.6) [79.6 %] | - | - |
| | | AD | 742 | 483 (65.1 %) | 483 (100 %) | 410 (55.3 %) | 74.7 (8.3) | 83.5 (6.4) [0.3 %] | - | 80.2 (8.8) [4.0 %] | 74.5 (8.2) [95.7 %] |
| **NACC - ADC4** | 827 | CN | 422 | 50 (11.8 %) | 15 (30.0 %) | 261 (61.8 %) | 79.4 (8.6) | 87.1 (7.8) [18.2 %] | 77.6 (7.8) [81.8 %] | - | - |
| | | AD | 405 | 156 (38.5 %) | 156 (100 %) | 217 (53.6 %) | 74.0 (7.3) | - | | 79.7 (12.4) [0.7 %] | 74.0 (7.3) [99.3 %] |
| **NACC - ADC5** | 989 | CN | 589 | 83 (14.1 %) | 16 (19.3 %) | 382 (64.9 %) | 81.9 (8.8) | 88.8 (6.7) [22.6 %] | 79.8 (8.2) [77.4 %] | - | - |
| | | AD | 400 | 150 (37.5 %) | 150 (100 %) | 215 (53.8 %) | 74.5 (7.9) | - | - | 76 (-) [0.2 %] | 74.5 (7.9) [99.8 %] |
| **NACC - ADC6** | 716 | CN | 357 | 39 (10.9 %) | 15 (38.5 %) | 240 (67.2 %) | 80.0 (8.9) | 87.0 (8.7) [21.0 %] | 78.1 (8.0) [79.0 %] | - | - |
| | | AD | 359 | 119 (33.1 %) | 119 (100 %) | 192 (53.5 %) | 74.4 (7.9) | - | - | 90 (-) [0.3 %] | 74.4 (7.9) [99.7 %] |
| **NACC - ADC7** | 1308 | CN | 772 | 39 (5.1 %) | 4 (10.3 %) | 497 (64.4 %) | 77.7 (7.8) | 84.0 (8.3) [9.3 %] | 77.1 (7.4) [90.7 %] | - | - |
| | | AD | 536 | 121 (22.6 %) | 121 (100 %) | 284 (53.0 %) | 72.9 (7.7) | - | - | - | 72.9 (7.7) [100 %] |
| **ADDNEURO** | 427 | CN | 182 | 0 (0.0%) | - | 102 (56.0 %) | 76.6 (6.3) | - | 76.6 (6.3) [100 %] | - | - |
| | | AD | 245 | 0 (0.0 %) | - | 158 (64.5 %) | 73.4 (6.2) | - | - | 78.2 (5.9) [8.6 %] | 73.0 (6.0) [91.4 %] |
| **ADNI** | 840 | CN | 344 | 1 (0.3 %) | 0 (0.0 %) | 172 (50.0 %) | 78.5 (6.8) | 84 (-) [0.3 %] | 78.5 (6.8) [99.7 %] | - | - |
| | | AD | 496 | 29 (5.8 %) | 29 (100 %) | 211 (42.5 %) | 75.5 (6.7) | - | - | 75.6 (6.7) [97.6 %] | 72.7 (7.5) [2.4 %] |
| **ADNI-DOD** | 79 | CN | 79 | 0 (0.0 %) | - | 0 (0.00 %) | 70.2 (5.3) | - | 70.2 (5.3) [100 %] | - | - |
| | | AD | - | - | - | - | - | - | - | - | - |
| **ADSP Extension** | 443 | CN | 160 | 26 (16.3 %) | 1 (3.8 %) | 93 (58.1 %) | 79.1 (7.5) | 82.7 (7.9) [5.6 %] | 78.8 (7.5) [94.4 %] | - | - |
| | | AD | 283 | 201 (71.0 %) | 201 (100 %) | 152 (53.7 %) | 74.2 (8.0) | - | - | 73 (-) [0.4 %] | 74.2 (8.0) [99.6 %] |
| **MAYO** | 1761 | CN | 1048 | 132 (12.6 %) | 0 (0.0 %) | 543 (51.8 %) | 74.2 (5.4) | 82.2 (5.0) [0.6 %] | 74.1 (5.3) [99.4 %] | - | - |
| | | AD | 713 | 227 (31.8 %) | 227 (100 %) | 414 (58.0 %) | 73.5 (5.1) | - | - | 73.5 (5.1) [99.4 %] | 68.8 (3.9) [0.6 %] |
| **MAYO2** | 230 | CN | 160 | 160 (100 %) | 0 (0.0 %) | 81 (50.6 %) | 83.8 (7.1) | 83.8 (7.1) [100 %] | - | - | - |
| | | AD | 70 | 70 (100 %) | 70 (100 %) | 48 (68.6 %) | 74.6 (5.7) | 84.4 (5.6) [98.6 %] | - | - | 85 (-) [1.4 %] |
| **MIRAGE** | 1098 | CN | 688 | 0 (0.0 %) | - | 403 (58.6 %) | 72.0 (7.2) | - | 72.0 (7.2) [100 %] | - | - |
| | | AD | 410 | 0 (0.0 %) | - | 254 (62.0 %) | 70.9 (6.5) | - | - | 74.5 (4.2) [1.0 %] | 70.9 (6.5) [99.0 %] |
| **MSBB** | 180 | CN | 31 | 31 (100 %) | 2 (6.5 %) | 23 (74.2 %) | 82.4 (7.6) | 82.4 (7.6) [100 %] | - | - | - |
| | | AD | 149 | 149 (100 %) | 149 (100 %) | 100 (67.1 %) | 78.1 (8.4) | 85.6 (7.1) [53.7 %] | - | - | 80.9 (9.0) [46.3 %] |
| **MTC** | 332 | CN | 113 | 0 (0.0 %) | - | 70 (61.9 %) | 73.1 (7.8) | - | 73.1 (7.8) [100 %] | - | - |
| | | AD | 219 | 1 (0.5 %) | - | 122 (55.7 %) | 74.4 (7.3) | 89 (-) [0.5 %] | - | 81.0 (8.3) [10.5 %] | 73.6 (6.8) [89.0 %] |
| **NIA-LOAD** | 3050 | CN | 1303 | 69 (5.3 %) | 6 (8.7 %) | 788 (60.5 %) | 74.5 (9.2) | 85.4 (7.0) [7.1 %] | 73.7 (8.8) [92.9 %] | - | - |
| | | AD | 1747 | 463 (26.5 %) | 463 (100 %) | 1145 (65.5 %) | 73.8 (6.8) | - | - | 84.2 (8.0) [0.5 %] | 73.7 (6.8) [99.5 %] |
| **OHSU** | 511 | CN | 326 | 326 (100 %) | 77 (23.6 %) | 171 (52.5 %) | 85.8 (7.3) | 85.8 (7.3) [100 %] | - | - | - |
| | | AD | 185 | 185 (100 %) | 185 (100 %) | 117 (63.2 %) | 85.3 (6.1) | - | - | 85.1 (6.5) [17.8 %] | 85.3 (6.0) [82.2 %] |

| | | | | | | | | | | | |
|---|---|---|---|---|---|---|---|---|---|---|---|
| **ROSMAP** | 1088 | CN | 528 | 291 (55.1 %) | 123 (42.3 %) | 383 (72.5 %) | 86.0 (7.2) | 88.1 (6.5) [58.5 %] | 83.1 (7.1) [41.5 %] | - | - |
| | | AD | 560 | 502 (89.6 %) | 502 (100 %) | 412 (73.6 %) | 84.4 (6.6) | - | - | 84.2 (6.5) [86.8 %] | 85.4 (7.0) [13.2 %] |
| **UM/VU/MSSM** | 1652 | CN | 931 | 42 (4.5 %) | 0 (0.0 %) | 594 (63.8 %) | 73.3 (7.7) | 80.9 (9.1) [6.3 %] | 72.8 (7.3) [93.7 %] | - | - |
| | | AD | 721 | 160 (22.2 %) | 160 (100 %) | 461 (63.9 %) | 74.3 (7.3) | 86.0 (7.1) [7.3 %] | - | 85.7 (6.1) [2.6 %] | 73.9 (7.1) [89.9 %] |
| **UPITT** | 2163 | CN | 870 | 0 (0.0 %) | - | 549 (63.1 %) | 75.4 (6.0) | - | 75.4 (6.0) [100 %] | - | - |
| | | AD | 1293 | 0 (0.0 %) | - | 822 (63.6 %) | 73.3 (6.6) | - | - | 78.1 (7.8) [7.3 %] | 72.9 (6.4) [92.7 %] |
| **WASHU** | 434 | CN | 139 | 2 (1.4 %) | 0 (0.0%) | 87 (62.6 %) | 76.9 (8.5) | 90.5 (0.7) [1.4 %] | 76.7 (8.4) [98.6 %] | - | - |
| | | AD | 295 | 28 (9.5 %) | 28 (100 %) | 171 (58.0 %) | 76.5 (8.8) | - | - | - | 76.5 (8.8) [100 %] |
| **WHICAP** | 622 | CN | 548 | 0 (0.0 %) | - | 330 (60.2 %) | 81.8 (6.7) | - | 81.8 (6.7) [100 %] | - | - |
| | | AD | 74 | 0 (0.0 %) | - | 53 (71.6 %) | 84.3 (7.7) | - | - | - | 84.3 (7.7) [100 %] |
| **TOTAL** | **25120** | CN | 12340 | 1699 (13.8 %) | 276 (16.2 %) | 7332 (59.4 %) | 78.3 (8.6) | 85.3 (7.1) [21.5 %] | 76.5 (8.0) [78.5 %] | - | - |
| | | AD | 12780 | 5027 (39.3 %) | 5027 (100 %) | 7582 (59.3 %) | 74.9 (7.8) | 85.1 (6.6) [1.8 %] | - | 77.8 (7.7) [16.2 %] | 74.3 (7.7) [82.0 %] |
| **REPLICATION SAMPLES** | | | | | | | | | | | |
| **ROTTERDAM** | 10150 | CN | 8824 | - | - | 4956 (56.2 %) | 77.5 (9.5) | 81.6 (8.8) [41.2 %] | 74.7 (8.9) [58.8 %] | - | - |
| | | AD | 1326 | - | - | 945 (71.3 %) | 83.7 (6.6) | - | - | 83.7 (6.6) [100 %] | - |
| **EADI** | 8571 | CN | 6502 | - | - | 3920 (60.3 %) | 80.4 (6.6) | 82.9 (6.3) [15.6 %] | 79.9 (6.7) [84.4 %] | - | - |
| | | AD | 2069 | - | - | 1359 (65.7 %) | 75.2 (8.2) | - | - | 75.2 (8.2) [100 %] | - |
| **EADB** | 21860 | CN | 12295 | - | - | 7155 (58.2 %) | 73.3 (8.0) | 79.7 (6.8) [0.2 %] | 73.3 (8.0) [99.8 %] | - | - |
| | | AD | 9565 | - | - | 6055 (63.3 %) | 74.9 (7.8) | - | - | - | 74.9 (7.8) [100 %] |

Subjects with a diagnosis of cognitively normal at their last examination prior to death were kept as controls, regardless of the presence of AD pathology.

**Table S4. Expanded sample demographics for Alzheimer's disease case-control association analyses in evaluated *APOE**4 carriers.**

| Cohort | | | | *APOE* *3/4 carriers | | | *APOE* *4/4 carriers | | |
|---|---|---|---|---|---|---|---|---|---|
| Name | Participants after QC (N) | Diagnosis (N) | | (N (%)) | Female (N (%)) | Age (Mean (SD)) | (N (%)) | Female (N (%)) | Age (Mean (SD)) |
| **DISCOVERY SAMPLES** | | | | | | | | | |
| **ACT** | 2269 | CN | 1645 | 299 (18.2 %) | 176 (58.9 %) | 82.7 (6.2) | 16 (1.0 %) | 11 (68.8 %) | 81.8 (6.8) |
| | | AD | 624 | 211 (33.8 %) | 128 (60.7 %) | 80.4 (5.9) | 40 (6.4 %) | 30 (75.0 %) | 74.7 (8.0) |
| **NACC - ADC1** | 2053 | CN | 490 | 99 (20.2 %) | 51 (51.5 %) | 77.2 (9.2) | 5 (1.0 %) | 4 (80.0 %) | 71.8 (3.3) |
| | | AD | 1563 | 765 (49.0 %) | 408 (53.3 %) | 73.0 (7.1) | 255 (16.3 %) | 118 (46.3 %) | 69.4 (5.9) |
| **NACC - ADC2** | 816 | CN | 125 | 23 (18.4 %) | 17 (73.9 %) | 76.1 (9.3) | 3 (2.4 %) | 2 (66.7 %) | 78.3 (8.5) |
| | | AD | 691 | 313 (45.3 %) | 173 (55.3 %) | 72.5 (6.3) | 113 (16.4 %) | 51 (45.1 %) | 68.8 (5.4) |
| **NACC - ADC3** | 1232 | CN | 490 | 94 (19.2 %) | 61 (64.9 %) | 76.0 (8.9) | 7 (1.4 %) | 4 (57.1 %) | 70.9 (3.0) |
| | | AD | 742 | 339 (45.7 %) | 182 (53.7 %) | 73.5 (7.2) | 96 (12.9 %) | 54 (56.3 %) | 70.8 (6.0) |
| **NACC - ADC4** | 827 | CN | 422 | 91 (21.6 %) | 54 (59.3 %) | 76.8 (8.4) | 11 (2.6 %) | 7 (63.6 %) | 77.0 (8.2) |
| | | AD | 405 | 166 (41.0 %) | 95 (57.2 %) | 72.7 (6.0) | 46 (11.4 %) | 24 (52.2 %) | 68.4 (4.8) |
| **NACC - ADC5** | 989 | CN | 589 | 105 (17.8 %) | 66 (62.9 %) | 79.9 (8.2) | 10 (1.7 %) | 6 (60.0 %) | 75.2 (6.6) |
| | | AD | 400 | 185 (46.3 %) | 108 (58.4 %) | 73.4 (6.7) | 48 (12.0 %) | 25 (52.1 %) | 68.6 (4.6) |
| **NACC - ADC6** | 716 | CN | 357 | 84 (23.5 %) | 55 (65.5 %) | 78.2 (8.3) | 5 (1.4 %) | 3 (60.0 %) | 72.4 (7.4) |
| | | AD | 359 | 156 (43.5 %) | 85 (54.5 %) | 72.5 (7.0) | 38 (10.6 %) | 17 (44.7 %) | 69.0 (5.4) |
| **NACC - ADC7** | 1308 | CN | 772 | 203 (26.3 %) | 125 (61.6 %) | 77.3 (7.7) | 14 (1.8 %) | 12 (85.7 %) | 74.1 (6.4) |
| | | AD | 536 | 239 (44.6 %) | 129 (54.0 %) | 72.3 (6.6) | 77 (14.4 %) | 39 (50.6 %) | 67.4 (6.7) |
| **ADDNEURO** | 427 | CN | 182 | 37 (20.3 %) | 22 (59.5 %) | 75.5 (6.4) | 3 (1.6 %) | 1 (33.3 %) | 73.3 (7.2) |
| | | AD | 245 | 102 (41.6 %) | 71 (69.6 %) | 72.8 (5.5) | 27 (11.0 %) | 19 (70.4 %) | 68.9 (4.9) |
| **ADNI** | 840 | CN | 344 | 82 (23.8 %) | 44 (53.7 %) | 76.4 (7.3) | 7 (2.0 %) | 2 (28.6 %) | 77.3 (4.5) |
| | | AD | 496 | 237 (47.8 %) | 102 (43.0 %) | 75.1 (6.1) | 83 (16.7 %) | 33 (39.8 %) | 72.1 (6.4) |
| **ADNI-DOD** | 79 | CN | 79 | 16 (20.3 %) | 0 (0.00 %) | 70.0 (6.2) | 2 (2.5 %) | 0 (0.00 %) | 72.5 (3.5) |
| | | AD | - | - | - | - | - | - | - |
| **ADSP Extension** | 443 | CN | 160 | 41 (25.6 %) | 22 (53.7 %) | 77.9 (6.9) | 1 (0.6 %) | 0 (0.00 %) | 77.0 (-) |
| | | AD | 283 | 134 (47.3 %) | 63 (47.0 %) | 72.6 (7.0) | 24 (8.5 %) | 11 (45.8 %) | 67.3 (6.3) |
| **MAYO** | 1761 | CN | 1048 | 250 (23.9 %) | 139 (55.6 %) | 73.9 (4.9) | 18 (1.7 %) | 9 (50.0 %) | 74.6 (2.7) |
| | | AD | 713 | 334 (46.8 %) | 192 (57.5 %) | 74.2 (4.7) | 123 (17.3 %) | 74 (60.2 %) | 71.4 (5.0) |
| **MAYO2** | 230 | CN | 160 | 31 (19.4 %) | 16 (51.6 %) | 82.3 (6.7) | 2 (1.3 %) | 1 (50.0 %) | 88.0 (2.8) |
| | | AD | 70 | 31 (44.3 %) | 23 (74.2 %) | 74.3 (5.6) | 5 (7.1 %) | 4 (80.0 %) | 67.8 (0.8) |
| **MIRAGE** | 1098 | CN | 688 | 210 (30.5 %) | 124 (59.0 %) | 71.4 (7.1) | 41 (6.0 %) | 28 (68.3 %) | 70.8 (6.9) |
| | | AD | 410 | 165 (40.2 %) | 107 (64.8 %) | 71.4 (6.2) | 63 (15.4 %) | 41 (65.1 %) | 68.3 (5.4) |
| **MSBB** | 180 | CN | 31 | 5 (16.1 %) | 5 (100 %) | 85.4 (3.6) | 0 (0.0 %) | - | - |
| | | AD | 149 | 56 (37.6 %) | 38 (67.9 %) | 77.4 (7.8) | 9 (6.0 %) | 5 (55.6 %) | 72.0 (7.4) |
| **MTC** | 332 | CN | 113 | 18 ( 15.9 %) | 13 (72.2 %) | 73.0 (6.1) | 1 (0.9 %) | 1 (100 %) | 73 (-) |
| | | AD | 219 | 99 (45.2 %) | 55 (55.5 %) | 73.6 (6.7) | 27 (12.3 %) | 10 (37.0 %) | 68.7 (5.6) |
| **NIA-LOAD** | 3050 | CN | 1303 | 412 (31.6 %) | 242 (58.7 %) | 72.0 (8.3) | 45 (3.5 %) | 26 (57.8 %) | 70.3 (8.00) |
| | | AD | 1747 | 948 (54.3 %) | 630 (66.5 %) | 73.0 (6.1) | 306 (17.5 %) | 183 (59.8 %) | 70.9 (6.4) |
| **OHSU** | 511 | CN | 326 | 60 (18.4 %) | 31 (51.7 %) | 84.4 (7.6) | 4 (1.2 %) | 1 (25.0 %) | 80.0 (5.6) |
| | | AD | 185 | 61 (33.0 %) | 33 (54.1 %) | 83.6 (6.3) | 6 (3.2 %) | 3 (50.0 %) | 81.3 (8.3) |

| | | | | | | | | | |
|---|---|---|---|---|---|---|---|---|---|
| **ROSMAP** | 1088 | CN | 528 | 82 (15.5 %) | 62 (75.6 %) | 84.3 (7.5) | 5 (1.0 %) | 5 (100 %) | 87.0 (4.6) |
| | | AD | 560 | 174 (31.1 %) | 128 (73.6 %) | 81.9 (6.7) | 14 (2.5 %) | 12 (85.7 %) | 78.4 (6.0) |
| **UM/VU/MSSM** | 1652 | CN | 931 | 190 (20.4 %) | 119 (62.6 %) | 72.1 (7.7) | 17 (1.8 %) | 11 (64.7 %) | 69.4 (5.1) |
| | | AD | 721 | 303 (42.0 %) | 201 (66.3 %) | 73.6 (6.6) | 101 (14.0 %) | 58 (57.4 %) | 69.7 (5.9) |
| **UPITT** | 2163 | CN | 870 | 144 (16.6 %) | 89 (61.8 %) | 73.4 (6.2) | 9 (1.0 %) | 5 (55.6 %) | 75.6 (2.3) |
| | | AD | 1293 | 586 (45.3 %) | 377 (64.3 %) | 72.5 (6.5) | 126 (9.7 %) | 69 (54.8 %) | 69.7 (5.8) |
| **WASHU** | 434 | CN | 139 | 29 (20.9 %) | 18 (62.1 %) | 72.7 (7.4) | 6 (4.3 %) | 3 (50.0 %) | 71.0 (5.8) |
| | | AD | 295 | 122 (41.4 %) | 73 (59.8 %) | 74.6 (7.7) | 24 (8.1 %) | 15 (62.5 %) | 70.9 (7.2) |
| **WHICAP** | 622 | CN | 548 | 102 (18.6 %) | 64 (62.7 %) | 80.9 (6.8) | 6 (1.1 %) | 3 (50.0 %) | 71.0 (5.8) |
| | | AD | 74 | 14 (18.9 %) | 10 (71.4 %) | 85.4 (5.9) | 1 (1.4 %) | 0 (0.00 %) | 70.9 (7.2) |
| **TOTAL** | **25120** | CN | 12340 | 2707 (21.9 %) | 1615 (59.7 %) | 76.2 (8.4) | 238 (1.9 %) | 145 (60.9 %) | 73.7 (7.4) |
| | | AD | 12780 | 5740 (44.9 %) | 3411 (59.4 %) | 73.8 (6.9) | 1652 (12.9 %) | 895 (54.2 %) | 70.1 (6.2) |
| **REPLICATION SAMPLES** | | | | | | | | | |
| **ROTTERDAM** | 10150 | CN | 8824 | 1868 (21.2 %) | 1034 (55.4 %) | 76.4 (9.3) | 150 (1.7 %) | 74 (49.3 %) | 73.8 (8.0) |
| | | AD | 1326 | 411 (31.0 %) | 282 (68.6 %) | 82.2 (6.4) | 86 (6.5 %) | 50 (58.1 %) | 78.2 (6.9) |
| **EADI** | 8571 | CN | 6502 | 1141 (17.5 %) | 672 (58.9 %) | 79.6 (6.4) | 62 (1.0 %) | 45 (72.6 %) | 77.8 (6.4) |
| | | AD | 2069 | 801 (38.7 %) | 529 (66.0 %) | 73.9 (7.4) | 205 (9.9 %) | 134 (65.4 %) | 69.4 (5.9) |
| **EADB** | 21860 | CN | 12295 | 2498 (20.3 %) | 1431 (57.3 %) | 72.4 (7.9) | 195 (1.6 %) | 107 (54.9 %) | 70.3 (7.5) |
| | | AD | 9565 | 3912 (40.9 %) | 2522 (64.5 %) | 74.0 (7.2) | 959 (10.0 %) | 551 (57.5 %) | 70.4 (6.8) |

**Table S5. Carrier frequencies of rs439401 across *APOE**2/3/4 genotypes, in the haplotype reference consortium[41], versus the current study discovery sample, using *APOE* filtering approach 1.** Note enrichment of *APOE**4-rs439401 haplotypes for *APOE**3/4 and 4/4 strata in the discovery sample compared to the haplotype reference consortium. Related individuals were filtered down to third degree relatedness, retaining a single sample per relatedness cluster.

**Haplotye Reference Consortum (HRC) r1.1**

| | rs439401 - T | | |
|---|---|---|---|
| **APOE** | **0** | **1** | **2** |
| **2/2** | 156 (100 %) | 0 (0.00 %) | 0 (0.00 %) |
| **2/3** | 1712 (54.09 %) | 1453 (45.91 %) | 0 (0.00 %) |
| **3/3** | 4864 (29.76 %) | 7840 (47.97 %) | 3639 (22.27 %) |
| **2/4** | 643 (99.69 %) | 2 (0.31 %) | 0 (0.00 %) |
| **3/4** | 3327 (53.44 %) | 2889 (46.40 %) | 10 (0.16 %) |
| **4/4** | 625 (99.21 %) | 5 (0.79 %) | 0 (0.00 %) |

**Discovery samples (including related individuals)**

| | rs439401 - T | | |
|---|---|---|---|
| **APOE** | **0** | **1** | **2** |
| **2/2** | 78 (97.50 %) | 1 (1.25 %) | 1 (1.25 %) |
| **2/3** | 1063 (52.31 %) | 962 (47.34 %) | 7 (0.34 %) |
| **3/3** | 3327 (27.67 %) | 6012 (50.00 %) | 2687 (22.34%) |
| **2/4** | 588 (97.84 %) | 13 (2.16 %) | 0 (0.00 %) |
| **3/4** | 4342 (51.48 %) | 4060 (48.13 %) | 33 (0.39 %) |
| **4/4** | 1856 (98.25 %) | 28 (1.48 %) | 5 (0.26 %) |

**Discovery samples (excluding related individuals)**

| | rs439401 - T | | |
|---|---|---|---|
| **APOE** | **0** | **1** | **2** |
| **2/2** | 75 (98.68 %) | 1 (1.32 %) | 0 (0.00 %) |
| **2/3** | 989 (52.36 %) | 894 (47.33 %) | 6 (0.32 %) |
| **3/3** | 3018 (27.42 %) | 5513 (50.10 %) | 2472 (22.47 %) |
| **2/4** | 508 (98.07 %) | 10 (1.93 %) | 0 (0.00 %) |
| **3/4** | 3825 (51.44 %) | 3584 (48.20 %) | 27 (0.36 %) |
| **4/4** | 1606 (98.35 %) | 22 (1.35 %) | 5 (0.31 %) |

**Table S6. Associations of rs439401 with case-control status in supporting *APOE**3/4 and 3/3 stratified analyses, using *APOE* filtering approach 1.**

| | Genotype distributions | | | AD Case-Control regression | |
|---|---|---|---|---|---|
| **Group/model** | **CN, carrier No. / Total No. (%)** | **AD, carrier No. / Total No. (%)** | **CN - AD, MAF (%)** | **OR (95% CI)** | **P-value** |
| **rs439401 - T allele tested** | | | | | |
| ***APOE* *3/4 - additive model** | | | | | |
| Discovery | 1320 / 2702 (48.9 %) | 2774 / 5734 (48.4 %) | 24.8 % - 24.3 % | 0.97 (0.88, 1.06) | 0.45 |
| ***APOE* *3/3 - additive model** | | | | | |
| Discovery | 5424 / 7519 (72.1 %) | 3276 / 4508 (72.7 %) | 47.4 % - 47.2 % | 1.00 (0.94, 1.05) | 0.86 |

**Table S7. Duplicate concordance across samples for rs439401.** Only genotype data that passed *APOE* filtering approach 1 and indicated European ancestry were used for these analyses (thus largely matching to samples considered in main analyses). In this subset, the total number of unique subjects with duplicates samples was N = 3804. Note there is subject overlap in the three respective analyses indicated in the table.

**Duplicates across SNP microarray samples (unique N = 1431)**

**Concordant**

| *APOE* | rs439401 - T: 0 | 1 | 2 |
|---|---|---|---|
| **2/2** | 6 | 0 | 0 |
| **2/3** | 72 | 62 | 0 |
| **3/3** | 182 | 340 | 167 |
| **2/4** | 27 | 0 | 0 |
| **3/4** | 240 | 223 | 1 |
| **4/4** | 106 | 4 | 0 |

**Discordant**

| *APOE* | rs439401 - T: 0 | 1 | 2 |
|---|---|---|---|
| **2/2** | 0 | 0 | 0 |
| **2/3** | 0 | 0 | 0 |
| **3/3** | 0 | 0 | 0 |
| **2/4** | 0 | 0 | 0 |
| **3/4** | 0 | 0 | 0 |
| **4/4** | 0 | 0 | 0 |

**Duplicates across NGS samples (unique N = 704)**

**Concordant**

| *APOE* | rs439401 - T: 0 | 1 | 2 |
|---|---|---|---|
| **2/2** | 0 | 0 | 0 |
| **2/3** | 0 | 1 | 0 |
| **3/3** | 12 | 11 | 3 |
| **2/4** | 1 | 0 | 0 |
| **3/4** | 5 | 7 | 0 |
| **4/4** | 0 | 0 | 0 |

**Discordant**

| *APOE* | rs439401 - T: 0 | 1 | 2 |
|---|---|---|---|
| **2/2** | 0 | 0 | 0 |
| **2/3** | 0 | 0 | 0 |
| **3/3** | 0 | 0 | 0 |
| **2/4** | 0 | 0 | 0 |
| **3/4** | 0 | 0 | 0 |
| **4/4** | 0 | 0 | 0 |

**Duplicates between SNP microarray and WGS samples (unique N = 2463)**

**Concordant**

| *APOE* | rs439401 - T: 0 | 1 | 2 |
|---|---|---|---|
| **2/2** | 6 | 0 | 0 |
| **2/3** | 114 | 118 | 1 |
| **3/3** | 365 | 647 | 307 |
| **2/4** | 53 | 1 | 0 |
| **3/4** | 368 | 344 | 4 |
| **4/4** | 131 | 1 | 0 |

**Discordant**

| *APOE* | rs439401 - T: 0 | 1 | 2 |
|---|---|---|---|
| **2/2** | 0 | 0 | 0 |
| **2/3** | 0 | 0 | 0 |
| **3/3** | 1* | 1* | 0 |
| **2/4** | 0 | 0 | 0 |
| **3/4** | 0 | 0 | 0 |
| **4/4** | 0 | 0 | 0 |

*Indicates 1 individual

**Table S8. Cohort and *APOE* genotype details of *APOE**4/4 subjects in the discovery carrying rs439401.** All shown participants were included in analyses using *APOE* filtering approach 1. Notably, one subject previously had *APOE**4/4 status called from the prior ADSP WES but now had a dubious call from the new ADSP WES data (first line in table), suggesting the subject is in fact an *APOE**3/4 carrier, which was also the provided and imputed *APOE* genotype.

| Cohort | rs439401_T | final *APOE* determined for approach 1 | prv_apoe (from demographics) | wgs_apoe (old ADSP extension + AMP-AD WGS) | wes_apoe (old ADSP discovery) | Subjects excluded in approach 2 | imp_apoe (TOPMed imputation) | rs429359_R2 (TOPMed imputation) | wgs_apoe (new ADSP WGS + AMP-AD WGS) | wes_apoe (new ADSP WES) | rs429359_DP (ADSP read depth) | rs7412_DP (ADSP read depth) | rs429358_AD (ADSP allelic distribution T,C) | rs7412_AD (ADSP allelic distribution C,T) |
|---|---|---|---|---|---|---|---|---|---|---|---|---|---|---|
| **CONTROLS** | | | | | | | | | | | | | | |
| ACT | 1 | 44 | 34 | | 44 | excluded | 34 | 0.92 | | ?4 | 11 | 3 | 1,10 | 3,0 |
| ADC1 | 1 | 44 | 44 | | | excluded | 34 | 0.93 | 34 | | 40 | 33 | 15,25 | 33,0 |
| ADC3 | 1 | 44 | 44 | | | included | 44 | 0.99 | | | | | | |
| ADC7 | 1 | 44 | 44 | | | included | 44 | 0.99 | 44 | | 48 | 38 | 0,48 | 38,0 |
| NIA-LOAD | 1 | 44 | 44 | | | excluded | 34 | 0.94 | | | | | | |
| MIRAGE | 2 | 44 | 44 | | | excluded | 33 | 0.90 | | | | | | |
| MIRAGE | 2 | 44 | 44 | | | excluded | 33 | 0.92 | | | | | | |
| MIRAGE | 1 | 44 | 44 | | | excluded | 34 | 0.92 | | | | | | |
| MIRAGE | 2 | 44 | 44 | | | excluded | 33 | 0.92 | | | | | | |
| MIRAGE | 1 | 44 | 44 | | | excluded | 34 | 0.92 | | | | | | |
| MIRAGE | 1 | 44 | 44 | | | excluded | 34 | 0.92 | | | | | | |
| MIRAGE | 1 | 44 | 44 | | | excluded | 33 | 0.92 | | | | | | |
| UPITT | 1 | 44 | 44 | | | excluded | 33 | 1 | | | | | | |
| WASHU | 1 | 44 | 44 | | | excluded | 33 | 0.93 | 33 | | 36 | 20 | 36,0 | 20,0 |
| **CASES** | | | | | | | | | | | | | | |
| ADC1 | 1 | 44 | 44 | | | included | 44 | 0.93 | | | | | | |
| ADC1 | 1 | 44 | 44 | | | excluded | 34 | 0.93 | | | | | | |
| ADC4 | 1 | 44 | 44 | | | included | 44 | 0.99 | | | | | | |
| ADC7 | 1 | 44 | 44 | | | included | 44 | 0.99 | 44 | | 66 | 35 | 0,66 | 35,0 |
| ADC7 | 1 | 44 | 44 | | | included | 44 | 0.99 | | | | | | |
| ADC7 | 1 | 44 | 44 | | | included | 44 | 9.99 | | | | | | |
| ADDNEURO | 1 | 44 | 44 | | | excluded | 33 | 0.98 | | | | | | |
| ADNI | 1 | 44 | 44 | 44 | | included | 44 | 0.99 | 44 | | 16 | 25 | 0,16 | 25,0 |
| NIA-LOAD | 1 | 44 | 44 | | | included | 44 | 0.94 | | | | | | |
| NIA-LOAD | 1 | 44 | 44 | | | included | 44 | 0.94 | | 44 | 24 | 9 | 0,24 | 9,0 |
| NIA-LOAD | 1 | 44 | 44 | | | included | 34 | 0.94 | | 44 | 28 | 6 | 0,28 | 6,0 |
| MAYO | 1 | 44 | 44 | | | included | 44 | 0.92 | | | | | | |
| MIRAGE | 1 | 44 | 44 | | | excluded | 34 | 0.92 | | | | | | |
| MIRAGE | 1 | 44 | 44 | | | excluded | 34 | 0.92 | | | | | | |
| MIRAGE | 1 | 44 | 44 | | | excluded | 34 | 0.92 | | | | | | |
| UPITT | 1 | 44 | 44 | | | excluded | 34 | 1 | | | | | | |
| UPITT | 2 | 44 | 44 | | | excluded | 33 | 1 | | | | | | |
| UM-VU-MSSM | 1 | 44 | 44 | | | included | 44 | 0.94 | | | | | | |
| UM-VU-MSSM | 2 | 44 | 44 | | | excluded | 33 | 0.98 | | | | | | |

**Table S9. Concordance rate between provided and TOPMed imputed *APOE**2/3/4 status across cohorts.** Note the rs7412 and rs429358 imputation (R2) scores for MIRAGE_370 and MIRAGE610. Very similar scores were observed for ADM_Q, ROSMAP_1B, and ROSMAP_1T, yet concordance rates between provided and imputed *APOE* were higher for those cohorts/arrays.

| Cohort/Array | Concordance rate provided versus imputed *APOE* | Participants with provided *APOE* (N) | Participants with imputed *APOE* (N) | Participants with provided and imputed *APOE* (N) | rs429358 $R^2$ | rs7412 $R^2$ |
|---|---|---|---|---|---|---|
| ACT | 95.37% | 2581 | 2709 | 2551 | 91.82% | 91.25% |
| ADC1 | 94.70% | 2731 | 2662 | 2662 | 93.28% | 89.79% |
| ADC2 | 95.23% | 926 | 924 | 922 | 93.19% | 90.89% |
| ADC3 | 99.34% | 1526 | 1517 | 1517 | 98.70% | 99.94% |
| ADC4 | 99.81% | 1051 | 1037 | 1034 | 98.72% | 99.96% |
| ADC5 | 99.59% | 1224 | 1223 | 1223 | 98.52% | 99.71% |
| ADC6 | 99.92% | 1333 | 1329 | 1329 | 98.73% | 99.98% |
| ADC7 | 99.72% | 1462 | 1454 | 1454 | 98.59% | 99.73% |
| ADM_O | 98.70% | 308 | 314 | 307 | 98.14% | 93.51% |
| ADM_Q | 88.45% | 278 | 328 | 277 | 88.33% | 80.21% |
| ADNI_1 | 93.13% | 757 | 742 | 742 | 93.03% | 87.97% |
| ADNI_DOD | 100.00% | 203 | 184 | 183 | 98.05% | 99.96% |
| ADNI_O25 | 99.37% | 812 | 796 | 796 | 99.13% | 99.55% |
| ADNI_OE | 100.00% | 360 | 349 | 348 | 98.54% | 99.98% |
| LOAD | 94.90% | 5220 | 5194 | 5194 | 93.79% | 90.51% |
| MAYO_1 | 93.59% | 2066 | 2046 | 2013 | 91.50% | 89.01% |
| MAYO_2 | 99.67% | 312 | 309 | 307 | 99.26% | 99.80% |
| MIRAGE_370 | 81.46% | 337 | 386 | 329 | 89.07% | 83.05% |
| MIRAGE_610 | 82.50% | 938 | 1102 | 937 | 91.99% | 83.24% |
| MTC | 98.78% | 409 | 530 | 409 | 98.56% | 99.79% |
| OHSU | 91.41% | 636 | 638 | 629 | 88.34% | 91.87% |
| ROSMAP_1B | 93.22% | 1120 | 1097 | 1091 | 90.45% | 80.38% |
| ROSMAP_1T | 94.13% | 581 | 580 | 579 | 91.39% | 82.10% |
| ROSMAP_2C | 98.69% | 381 | 382 | 381 | 98.06% | 99.98% |
| ROSMAP_3BU | 98.20% | 424 | 452 | 389 | 98.88% | 99.93% |
| UPITT | 96.82% | 2436 | 2363 | 2359 | 99.53% | 99.61% |
| UVM_A | 95.89% | 1147 | 1150 | 1144 | 94.10% | 91.39% |
| UVM_B | 95.22% | 857 | 864 | 857 | 98.03% | 99.98% |
| WASHU | 92.22% | 668 | 670 | 668 | 92.93% | 92.63% |
| WHICAP | 97.20% | 642 | 647 | 642 | 98.35% | 97.22% |

**Table S10. Discordance rate between *APOE* genotypes from different sources.** Discordance rates in *APOE**4/4 carriers was determined only for provided *APOE**4/4 subjects.

**Full sample**

discordance rate = 4.3%

| | imputed *APOE* | | | | | |
|---|---|---|---|---|---|---|
| provided *APOE* | 2/2 | 2/3 | 2/4 | 3/3 | 3/4 | 4/4 |
| 2/2 | 68 | 5 | 1 | 2 | 2 | 0 |
| 2/3 | 10 | 1859 | 14 | 108 | 18 | 1 |
| 2/4 | 0 | 21 | 521 | 7 | 31 | 0 |
| 3/3 | 0 | 80 | 5 | 11501 | 182 | 13 |
| 3/4 | 0 | 23 | 11 | 352 | 7837 | 32 |
| 4/4 | 0 | 0 | 1 | 13 | 135 | 1708 |

discordance rate = 2.8%

| | imputed *APOE* | | | | | |
|---|---|---|---|---|---|---|
| wgs *APOE* | 2/2 | 2/3 | 2/4 | 3/3 | 3/4 | 4/4 |
| 2/2 | 8 | 1 | 0 | 0 | 0 | 0 |
| 2/3 | 0 | 214 | 0 | 19 | 1 | 0 |
| 2/4 | 0 | 0 | 43 | 1 | 4 | 0 |
| 3/3 | 0 | 4 | 0 | 1322 | 9 | 0 |
| 3/4 | 0 | 1 | 2 | 24 | 696 | 1 |
| 4/4 | 0 | 0 | 0 | 0 | 2 | 115 |

discordance rate = 0.9%

| | provided *APOE* | | | | | |
|---|---|---|---|---|---|---|
| wgs *APOE* | 2/2 | 2/3 | 2/4 | 3/3 | 3/4 | 4/4 |
| 2/2 | 11 | 0 | 0 | 0 | 0 | 0 |
| 2/3 | 0 | 256 | 0 | 0 | 0 | 0 |
| 2/4 | 0 | 0 | 61 | 0 | 2 | 0 |
| 3/3 | 0 | 2 | 1 | 1538 | 10 | 0 |
| 3/4 | 0 | 2 | 1 | 6 | 883 | 0 |
| 4/4 | 0 | 0 | 0 | 0 | 1 | 138 |

***APOE*** ***4/4 sample (determined through *APOE* filtering approach 1)**

discordance rate = 7.9%

| | imputed *APOE* | | | | | |
|---|---|---|---|---|---|---|
| provided *APOE* | 2/2 | 2/3 | 2/4 | 3/3 | 3/4 | 4/4 |
| 2/2 | - | - | - | - | - | - |
| 2/3 | - | - | - | - | - | - |
| 2/4 | - | - | - | - | - | - |
| 3/3 | - | - | 0 | 0 | 0 | 1 |
| 3/4 | - | - | 0 | 0 | 1 | 8 |
| 4/4 | - | - | 1 | 12 | 134 | 1708 |

discordance rate = 1.7%

| | imputed *APOE* | | | | | |
|---|---|---|---|---|---|---|
| wgs *APOE* | 2/2 | 2/3 | 2/4 | 3/3 | 3/4 | 4/4 |
| 2/2 | - | - | - | - | - | - |
| 2/3 | - | - | - | - | - | - |
| 2/4 | - | - | - | - | - | - |
| 3/3 | - | - | - | - | - | - |
| 3/4 | - | - | - | - | - | - |
| 4/4 | - | - | 0 | 0 | 2 | 115 |

discordance rate = 0.7%

| | provided *APOE* | | | | | |
|---|---|---|---|---|---|---|
| wgs *APOE* | 2/2 | 2/3 | 2/4 | 3/3 | 3/4 | 4/4 |
| 2/2 | - | - | - | - | - | - |
| 2/3 | - | - | - | - | - | - |
| 2/4 | - | - | - | - | - | - |
| 3/3 | - | - | - | - | - | - |
| 3/4 | - | - | - | - | - | - |
| 4/4 | - | - | - | 0 | 1 | 138 |

***APOE*** ***4/4 cases sample (determined through *APOE* filtering approach 1)**

discordance rate = 6.8%

| | imputed *APOE* | | | | | |
|---|---|---|---|---|---|---|
| provided *APOE* | 2/2 | 2/3 | 2/4 | 3/3 | 3/4 | 4/4 |
| 2/2 | - | - | - | - | - | - |
| 2/3 | - | - | - | - | - | - |
| 2/4 | - | - | - | - | - | - |
| 3/3 | - | - | 0 | 0 | 0 | 1 |
| 3/4 | - | - | 0 | 0 | 0 | 7 |
| 4/4 | - | - | 1 | 5 | 104 | 1511 |

discordance rate = 1.9%

| | imputed *APOE* | | | | | |
|---|---|---|---|---|---|---|
| wgs *APOE* | 2/2 | 2/3 | 2/4 | 3/3 | 3/4 | 4/4 |
| 2/2 | - | - | - | - | - | - |
| 2/3 | - | - | - | - | - | - |
| 2/4 | - | - | - | - | - | - |
| 3/3 | - | - | - | - | - | - |
| 3/4 | - | - | - | - | - | - |
| 4/4 | - | - | 0 | 0 | 14 | 146 |

discordance rate = 0.8%

| | provided *APOE* | | | | | |
|---|---|---|---|---|---|---|
| wgs *APOE* | 2/2 | 2/3 | 2/4 | 3/3 | 3/4 | 4/4 |
| 2/2 | - | - | - | - | - | - |
| 2/3 | - | - | - | - | - | - |
| 2/4 | - | - | - | - | - | - |
| 3/3 | - | - | - | - | - | - |
| 3/4 | - | - | - | - | - | - |
| 4/4 | - | - | - | 0 | 1 | 128 |

***APOE*** ***4/4 controls sample (determined through *APOE* filtering approach 1)**

discordance rate = 15.8%

| | imputed *APOE* | | | | | |
|---|---|---|---|---|---|---|
| provided *APOE* | 2/2 | 2/3 | 2/4 | 3/3 | 3/4 | 4/4 |
| 2/2 | - | - | - | - | - | - |
| 2/3 | - | - | - | - | - | - |
| 2/4 | - | - | - | - | - | - |
| 3/3 | - | - | - | - | - | - |
| 3/4 | - | - | - | 0 | 1 | 1 |
| 4/4 | - | - | - | 7 | 30 | 197 |

discordance rate = 0%

| | imputed *APOE* | | | | | |
|---|---|---|---|---|---|---|
| wgs *APOE* | 2/2 | 2/3 | 2/4 | 3/3 | 3/4 | 4/4 |
| 2/2 | - | - | - | - | - | - |
| 2/3 | - | - | - | - | - | - |
| 2/4 | - | - | - | - | - | - |
| 3/3 | - | - | - | - | - | - |
| 3/4 | - | - | - | - | - | - |
| 4/4 | - | - | - | 0 | 0 | 9 |

discordance rate = 0%

| | provided *APOE* | | | | | |
|---|---|---|---|---|---|---|
| wgs *APOE* | 2/2 | 2/3 | 2/4 | 3/3 | 3/4 | 4/4 |
| 2/2 | - | - | - | - | - | - |
| 2/3 | - | - | - | - | - | - |
| 2/4 | - | - | - | - | - | - |
| 3/3 | - | - | - | - | - | - |
| 3/4 | - | - | - | - | - | - |
| 4/4 | - | - | - | - | 0 | 10 |

***APOE*** ***4/4-rs439401 haplotype cases sample (determined through *APOE* filtering approach 1)**

discordance rate = 47.4%

| | imputed *APOE* | | | | | |
|---|---|---|---|---|---|---|
| provided *APOE* | 2/2 | 2/3 | 2/4 | 3/3 | 3/4 | 4/4 |
| 2/2 | - | - | - | - | - | - |
| 2/3 | - | - | - | - | - | - |
| 2/4 | - | - | - | - | - | - |
| 3/3 | - | - | - | - | - | - |
| 3/4 | - | - | - | - | - | - |
| 4/4 | - | - | - | 3 | 6 | 10 |

discordance rate = 25%

| | imputed *APOE* | | | | | |
|---|---|---|---|---|---|---|
| wgs *APOE* | 2/2 | 2/3 | 2/4 | 3/3 | 3/4 | 4/4 |
| 2/2 | - | - | - | - | - | - |
| 2/3 | - | - | - | - | - | - |
| 2/4 | - | - | - | - | - | - |
| 3/3 | - | - | - | - | - | - |
| 3/4 | - | - | - | - | - | - |
| 4/4 | - | - | 0 | 0 | 1 | 3 |

discordance rate = 0%

| | provided *APOE* | | | | | |
|---|---|---|---|---|---|---|
| wgs *APOE* | 2/2 | 2/3 | 2/4 | 3/3 | 3/4 | 4/4 |
| 2/2 | - | - | - | - | - | - |
| 2/3 | - | - | - | - | - | - |
| 2/4 | - | - | - | - | - | - |
| 3/3 | - | - | - | - | - | - |
| 3/4 | - | - | - | - | - | - |
| 4/4 | - | - | - | - | - | 4 |

***APOE*** ***4/4-rs439401 haplotype controls sample (determined through *APOE* filtering approach 1)**

discordance rate = 84.6%

| | imputed *APOE* | | | | | |
|---|---|---|---|---|---|---|
| provided *APOE* | 2/2 | 2/3 | 2/4 | 3/3 | 3/4 | 4/4 |
| 2/2 | - | - | - | - | - | - |
| 2/3 | - | - | - | - | - | - |
| 2/4 | - | - | - | - | - | - |
| 3/3 | - | - | - | - | - | - |
| 3/4 | - | - | - | 0 | 1 | 0 |
| 4/4 | - | - | - | 6 | 5 | 2 |

discordance rate = -

| | imputed *APOE* | | | | | |
|---|---|---|---|---|---|---|
| wgs *APOE* | 2/2 | 2/3 | 2/4 | 3/3 | 3/4 | 4/4 |
| 2/2 | - | - | - | - | - | - |
| 2/3 | - | - | - | - | - | - |
| 2/4 | - | - | - | - | - | - |
| 3/3 | - | - | - | - | - | - |
| 3/4 | - | - | - | - | - | - |
| 4/4 | - | - | - | - | - | - |

discordance rate = -

| | provided *APOE* | | | | | |
|---|---|---|---|---|---|---|
| wgs *APOE* | 2/2 | 2/3 | 2/4 | 3/3 | 3/4 | 4/4 |
| 2/2 | - | - | - | - | - | - |
| 2/3 | - | - | - | - | - | - |
| 2/4 | - | - | - | - | - | - |
| 3/3 | - | - | - | - | - | - |
| 3/4 | - | - | - | - | - | - |
| 4/4 | - | - | - | - | - | - |

9. American Psychiatric Association. Diagnostic and Statistical Manual of Mental Disorders 5th edn. *Am Psychiatr Assoc*. Published online 2013.

10. American Psychiatric Association. Diagnostic and Statistical Manual of Mental Disorders 4th edn. *Am Psychiatr Assoc*. Published online 1994.

11. Hughes CP, Berg L, Danziger WL, Coben LA, Martin RL. A new clinical scale for the staging of dementia. *Br J Psychiatry*. 1982;140(6):566-572. doi:10.1192/bjp.140.6.566

12. Braak H, Braak E. Neuropathological stageing of Alzheimer-related changes. *Acta Neuropathol*. 1991;82:239-259. doi:10.1109/ICINIS.2015.10

13. Mirra SS, Heyman A, McKeel D, et al. The Consortium to Establish a Registry for Alzheimer's Disease (CERAD). Part II. Standardization of the neuropathologic assessement of Alzheimer's disease. *Neurology*. 1991;41:479-486. doi:10.1017/CBO9781107415324.004

14. Hyman BT, Trojanowski JQ. Editorial on Consensus Recommendations for the Postmortem Diagnosis of Alzheimer Disease from the National Institute on Aging and the Reagan Institute

Working Group on Diagnositic Criteria for the Neuropathological Assessment of Alzheimr Disease. *J Neuropathol Exp Neurol*. 1997;56(10):1095-1097.

26. Bennett DA, Wilson RS, Schneider JA, et al. Natural history of mild cognitive impairment in older

persons. *Neurology*. 2002;59(2):198-205. doi:10.1212/WNL.59.2.198

37. Zhu C, Farrell J, Kuzma A, Schellenberg G, Farrer L. Precision genotyping of APOE from whole genome sequencing. In: *American Society of Human Genetics Congress*. ; 2019.

38. Naj AC, Jun G, Beecham GW, et al. Common variants at MS4A4/MS4A6E, CD2AP, CD33 and EPHA1 are associated with late-onset Alzheimer’s disease. *Nat Genet*. 2011;43(5):436-443. doi:10.1038/ng.801

39. Wattmo C, Londos E, Minthon L. Risk factors that affect life expectancy in alzheimer’s disease: A 15-year follow-up. *Dement Geriatr Cogn Disord*. 2014;38(5-6):286-299. doi:10.1159/000362926

40. Kuzma A, Valladares O, Cweibel R, et al. NIAGADS: The NIA Genetics of Alzheimer’s Disease Data Storage Site. *Alzheimer’s Dement*. 2016;12(11):1200-1203. doi:10.1016/j.jalz.2016.08.018

41. McCarthy S, Das S, Kretzschmar W, et al. A reference panel of 64,976 haplotypes for genotype imputation. *Nat Genet*. 2016;48(10):1279-1283. doi:10.1038/ng.3643
